# The Real-World Costs of GLP-1 Receptor Agonist Treatment

**DOI:** 10.1101/2025.10.24.25338255

**Authors:** David Wennberg, Henriette Coetzer, Amy Marr, Caroline Margiotta, Razia Hashmi, Jason Kay, Vikrant Vats, Stuart Hagen, Laura Anatale-Tardiff, Sandhya Rao, Mark Mugiishi, Jonathan Skinner

## Abstract

**Background:** The high cost and widespread use of glucagon-like peptide 1 receptor agonists (GLP-1RAs) are a concern for healthcare budgets. Whether GLP-1RA use reduces other health care spending is unclear.

**Methods:** We conducted a cohort study using insurance claims data for United States adults aged 18-64 from 2016-2024, matching GLP-1RA treated members with untreated members (controls) on baseline demographics, clinical conditions, hospitalization, and medication use. Primary outcomes were per member per month (PMPM) healthcare costs overall and by service type, analyzed separately for members with and without diabetes.

**Results:** Among 742,824 matched treated and control members, 55.6% had diabetes. In year 1 following GLP-1RA initiation, total costs were 68.7% higher in treated members (95% CI, 68.0%-69.4%, $743 PMPM difference); in years 2-6 costs were 44.8% higher (95% CI, 43.7%-45.9%; $530 PMPM difference). Excluding GLP-1RA costs, treated members had 5.8% higher costs in year 1 (95% CI 5.1%-6.5%) and 4.1% higher costs (95% CI 3.0% - 5.2%) in years 2-6. Among treated members with diabetes, cost increases were modest: 3.8% (95% CI 2.8% - 4.8%) in year 1 and 0.8% in years 2-6 (95% CI 0.6%-2.2%), with non-GLP-1RA pharmacy and provider visits offset by reduced admissions and dialysis. Treated members without diabetes had more substantial cost increases: 8.9% in year 1 (95% CI 7.7% - 10.1%) and 9.7% in years 2-6 (95% CI 8.0% - 11.4%).

**Conclusions:** GLP-1RA treatment was associated with increases in spending on healthcare net of the GLP-1RA cost, particularly in members without diabetes.

**Key Points:** *Question:* What are the real-world costs of GLP-1 receptor agonist (GLP-1RA) treatment for adults with diabetes and other conditions?

*Findings:* GLP-1RAs treatment is associated with substantially increased healthcare costs. Excluding the costs of GLP-1RAs, treated adults with diabetes have modest increases in costs; however, those treated without diabetes have costs that are.9% higher than those not receiving the drug.

*Meaning:* Medical costs of using GLP-1RAs for those without diabetes go far beyond the costs of the agents. An estimated 40% of the US adult population are eligible for treatment for obesity. Treating them with GLP-1RAs would have a substantial impact on insurance costs.

## Introduction

GLP-1 receptor agonist medications (GLP-1RAs) mimic the natural hormone GLP-1 by regulating blood sugar and suppressing appetite, providing significant clinical benefits for the management of diabetes and obesity.^1^ For people with diabetes, GLP-1RAs can lower risk of cardiovascular events, improve lipid profiles, reduce fatty liver disease, lower dementia risk, reduce sleep apnea, and delay progression of diabetes-related nephropathy.^2-7^ They also promote weight loss of between 4% and 20% of body weight over a year.^2,8-11^ However, monthly costs of $900-$1300 resulted in GLP-1RAs accounting for over 13% of insurance premium costs in 2023, prompting insurance coverage restrictions. ^12,13^

Prior studies suggest that the cost of the drugs can be partially offset by improvements in health and associated lower medical care, and for those treated for obesity, by reduced risk of diabetes and associated complications.^14-19^ These studies have accounted for the direct GLP-1RA medication and related visit costs or the rare adverse side effects, but data on the real-world costs of other medical services remain limited.^20,21^

Using a large, national, commercial insurance database, we followed people starting GLP-1RAs and a matched control population for up to 6 years. We evaluated the healthcare costs associated with GLP-1RA treatment for all treated and the costs for those with and without diabetes. We were interested in overall costs and costs for specific types of services among treated versus controls.

## Methods

### Ethics statement

This retrospective cohort study was exempted from full review by the Sterling Institutional Review Board (Protocol #12167) and granted a waiver of informed consent.

### Setting and Data Sources

This study used the National Data Warehouse, managed by Blue Cross Blue Shield Association, housing BCBS Plan submitted data (e.g., medical claims, membership data). Adults aged 18-64 between 1 January 2016 and 31 August 2024 with commercial insurance, residing in the United States, with primary medical and pharmacy benefits, were potentially eligible for the study (N=24,987,055, Table S1).

### Cohort Construction

#### Members receiving GLP-1RAs (treated)

We identified 1,725,097 members with ≥1 pharmacy claim for a GLP-1RA product (tirzepatide, albiglutide, semaglutide, liraglutide, lixisenatide, dulaglutide, or exenatide) between 1 January 2017 and 29 February 2024 who had ≥12 months of continuous enrollment prior to their first GLP-1RA prescription (baseline).

Member were classified as having a condition if they had ≥1 inpatient facility claim with relevant ICD10 diagnoses or ≥2 ambulatory facility or professional claims (excluding claims for ambulances, DME, labs, and imaging) on separate days in their baseline year.

Members were excluded if in their baseline they had: contraindications for GLP-1RAs (e.g., pregnancy), high-cost conditions (e.g. cancers except squamous and basal cell carcinoma, mechanical ventilation >96 hours), conditions making long term GLP-1RA use difficult (e.g., dementia), or bariatric procedures. We required all members to have ≥6 months follow-up post first GLP-1RA prescription. These resulted in 936,777 eligible treated members for the study (Table S1).

#### Members not receiving GLP-1RAs (controls)

Eligible control members had no prescription fill for a GLP-1RA product between 1 January 2017 and 29 February 2024 and at least 18 months of continuous eligibility (N= 23,261,958). Eligible control members were randomly assigned a date within a 3-month period (quarters) from 1 January 2017 through 29 February 2024 defined as a proxy exposure date (pseudo-index date).^22^ Members aged 18 to 64 years as of their pseudo-index date and with >12 months of continuous eligibility in the baseline period and ≥6 months eligibility in the follow-up period were identified (N=22,716,175). The same baseline exclusion requirements as treated members were applied resulting in 18,281,409 eligible controls for the study.

#### Matching

Eligible control members were exact-matched to treated members on gender, age group (18-24, then 5-year increments to 64), year-quarter of the index date or pseudo-index date, and baseline prediabetes, type 1 diabetes mellitus, type 2 diabetes mellitus with or without complications, obesity, and overweight. Eligible control members were propensity score matched using a caliper of 0.05 standard deviations of the logit of the propensity score to treated members on baseline anxiety, depression, asthma, COPD, obstructive sleep apnea, hypertension, dyslipidemia, cerebrovascular disease, chronic kidney disease, coronary artery disease, heart failure, early liver disease (e.g., non-alcoholic steatohepatitis), late stage liver disease (e.g., cirrhosis), alcohol or opiate use disorder, pancreatitis, psoriatic and rheumatic disease, and weight-bearing joint pain. Eligible control members were also propensity score-matched on their use of non-GLP-1RA diabetes medications (insulin, SGLT2 inhibitors, sulfonylureas, meglitinide analogues, thiazolidinediones, DPP-4 Inhibitors, and biguanides), cardiovascular medications (loop diuretics, carbonic anhydrase inhibitors, thiazides, ACE inhibitors, ARBs, or neprilysin inhibitors), number of inpatient stays, and number of unique drug classes in their baseline year. Finally, eligible members were propensity score matched on race and ethnicity using the RAND Bayesian Improved First Name Surname Geocoding race/ethnicity imputation.^23^ Matching resulted in 742,824 treated and control members respectively. The reduction in eligible treated is primarily a result of a lower use of SGLT2 and DPP-4 inhibitors among eligible controls (Table S2).

### Outcomes

We calculated expenditures for medications and medical care per member per month (PMPM) based on the allowed amount. PMPM rates were calculated for overall costs and several sub-categories of healthcare services (e.g., provider visits). To reduce the impact of outliers, we applied a $500,000 annualized cap on total medical and pharmacy costs at the member level, scaling all component costs for members exceeding that level. We performed two additional subgroup analyses: 1) we stratified the results by later generation GLP-1RAs (semaglutide and tirzepatide) versus older generation (all others); and 2) we looked at differences in costs for claims with gastrointestinal conditions diagnoses (e.g. nausea and vomiting), the most common side effects of GLP-1RAs. The medical care component of the consumer price index (BLS series CUUR0000SAM) was used to adjust costs to August 2024 dollars.^24^

### Follow up

Members were followed until the earliest of disenrollment from medical or pharmacy benefits or the end of the study period. Median follow-up for treated was 20 months [IQR 14-30 months] and for controls 19 months [IQR 13-28 months]. GLP-1RA persistence was assessed by aggregating each member’s GLP-1RA treatment periods, allowing gaps up to 90 days. Treatment persistence was generally short: 52.9% of those with diabetes and 66.5% of those without diabetes were treated for <1 year, 29.1% of those with diabetes and 27.5% of those without diabetes were treated for 1-<2 years, and 18.5% of those with diabetes and 6.0% of those without diabetes were treated for ≥2 years.

### Statistical Analysis

Analyses were conducted with R Studio Version 2024.04.2. Baseline characteristics, and healthcare costs and utilization were summarized using mean values and 95% confidence intervals for continuous variables and frequency distributions for categorical variables. Statistical significance of differences between cohorts in mean costs was assessed via Student’s T-Tests and differences in proportions via Chi-square tests.

## Results

Baseline characteristics of treated (N=742,824) and controls (N=742,824) across demographics, conditions, medications, and inpatient stays were well-balanced (Table 1). Treated and controls had a mean age of 50 years and were more likely to be female (60.8%). 55.6% of members had diabetes, and a majority had 1 or more other chronic conditions. Members without diabetes had higher rates of obesity than those with diabetes and comparable rates of obstructive sleep apnea.

**Table 1:** Characteristics of Matched GLP-1RA User and Control Cohorts.

| Characteristic considered in the matching algorithm | Full Matched Population |  |  | Members with Diabetes |  |  | Members without Diabetes |  |  |
| --- | --- | --- | --- | --- | --- | --- | --- | --- | --- |
|  | Treated | Controls | SMD | Treated | Controls | SMD | Treated | Controls | SMD |
|  | N =<br>742,824 | N =<br>742,824 |  | N =<br>413,404 | N =<br>413,404 |  | N =<br>329,420 | N =<br>329,420 |  |
| Age at Index (Years), mean (SD) | 50 (10) | 50 (10) | 0.01 | 52.4 (8.8) | 52.5 (8.9) | 0.01 | 46.7 (10.2) | 46.7 (10.3) | 0.01 |
| Gender (% Female) | 60.8% | 60.8% | 0 | 49.0% | 49.0% | 0 | 75.7% | 75.7% | 0 |
| Race/Ethnicity |  |  |  |  |  |  |  |  |  |
| White | 66.7% | 65.5% | 0.03 | 63.5% | 61.1% | 0.05 | 70.5% | 70.9% | 0.01 |
| Black | 7.8% | 8.1% | 0.01 | 8.3% | 8.8% | 0.02 | 7.2% | 7.2% | 0 |
| Hispanic | 9.1% | 9.7% | 0.02 | 10.7% | 11.7% | 0.03 | 7.1% | 7.1% | 0 |
| Other | 3.8% | 4.0% | 0.01 | 4.6% | 5.1% | 0.02 | 2.9% | 2.5% | 0.02 |
| Unknown | 12.6% | 12.9% | 0.01 | 12.8% | 13.3% | 0.01 | 12.3% | 12.3% | 0 |
| Type 1 Diabetes | 1.1% | 1.1% | 0 | 2.0% | 2.0% | 0 | 0.0% | 0.0% | 0 |
| Type 2 Diabetes with complications | 19.1% | 19.1% | 0 | 34.3% | 34.3% | 0 | 0.0% | 0.0% | 0 |
| Type 2 Diabetes without complications | 35.4% | 35.4% | 0 | 63.7% | 63.7% | 0 | 0.0% | 0.0% | 0 |
| Obesity | 39.9% | 39.9% | 0 | 35.7% | 35.7% | 0 | 45.1% | 45.1% | 0 |
| Liver Disease | 3.5% | 3.3% | 0.01 | 4.2% | 4.0% | 0.01 | 2.7% | 2.2% | 0.01 |
| Hypertension | 50.1% | 51.3% | 0.02 | 61.8% | 63.6% | 0.04 | 35.4% | 35.9% | 0.01 |
| Anxiety | 15.5% | 15.6% | 0 | 11.7% | 11.9% | 0.01 | 20.1% | 20.2% | 0.00 |
| Depression | 11.4% | 11.3% | 0 | 9.4% | 9.4% | 0.00 | 14.0% | 13.8% | 0.01 |
| Heart Failure | 1.4% | 1.3% | 0.01 | 2.1% | 2.1% | 0.00 | 0.6% | 0.3% | 0.03 |
| Coronary Artery Disease | 4.8% | 4.6% | 0.01 | 6.9% | 6.6% | 0.01 | 2.1% | 1.9% | 0.01 |
| Chronic Kidney Disease | 2.8% | 2.7% | 0 | 4.2% | 4.2% | 0 | 0.9% | 0.7% | 0.02 |
| Obstructive Sleep Apnea | 10.4% | 9.8% | 0.02 | 11.0% | 10.3% | 0.02 | 9.7% | 9.1% | 0.02 |
| Prior Treatment |  |  |  |  |  |  |  |  |  |
| Insulin | 7.6% | 7.2% | 0.02 | 13.6% | 12.9% | 0.02 | 0.0% | 0.0% | 0.00 |
| SGLT2 Inhibitors | 10.3% | 9.0% | 0.05 | 18.6% | 16.2% | 0.06 | 0.0% | 0.0% | 0.00 |
| Sulfonylureas<sup>a</sup> | 12.2% | 11.9% | 0.01 | 21.9% | 21.3% | 0.01 | 0.0% | 0.0% | 0.00 |
| DPP-4 Inhibitors | 5.9% | 5.5% | 0.01 | 10.5% | 9.9% | 0.02 | 0.0% | 0.0% | 0.00 |
| Biguanides | 41.3% | 41.6% | 0.01 | 72.4% | 73.8% | 0.03 | 2.3% | 1.3% | 0.08 |
| Number of Non-Bariatric Inpatient Stays |  |  |  |  |  |  |  |  |  |
| 0 | 96.1% | 96.4% | 0.01 | 95.0% | 95.2% | 0.01 | 97.6% | 97.8% | 0.01 |
| 1 | 3.2% | 3.0% | 0.01 | 4.2% | 3.9% | 0.01 | 2.0% | 1.9% | 0.01 |
| 2+ | 0.6% | 0.6% | 0 | 0.9% | 0.9% | 0 | 0.3% | 0.3% | 0.01 |
| Number of other prescription classes used at baseline |  |  |  |  |  |  |  |  |  |
| 0 | 4.9% | 4.7% | 0.01 | 5.2% | 4.7% | 0.02 | 4.5% | 4.6% | 0 |
| 1 to 3 | 28.0% | 27.6% | 0.01 | 30.4% | 29.8% | 0.01 | 25.0% | 24.9% | 0 |
| 4 to 6 | 28.7% | 29.0% | 0.01 | 29.3% | 29.7% | 0.01 | 28.0% | 28.1% | 0 |
| 7 to 9 | 18.9% | 19.1% | 0.01 | 17.9% | 18.3% | 0.01 | 20.1% | 20.2% | 0 |
| 10+ | 19.5% | 19.6% | 0 | 17.1% | 17.5% | 0.01 | 22.4% | 22.3% | 0 |

<sup>a</sup>The sulfonylureas category also includes meglitinide analogues and thiazolidinediones.

Matching on clinical conditions, pharmacy use, and inpatient stay counts resulted in baseline total health care costs that were slightly higher in controls: 2.3% (95% CI: 1.7%–2.9%; p < 0.001) higher in the overall population, 2.4% (95% CI: 1.6%–3.2%; p < 0.001) higher in controls with diabetes and 2.2% (95% CI: 1.2%–3.1%; p < 0.001) higher in controls without diabetes (Figure 1 and Tables 2 and 3). While baseline costs between control and treated groups varied across subcategories of care, the absolute differences were small. Utilization rates in treated and controls reflected the cost differences (Table S3).

**Table 2:**
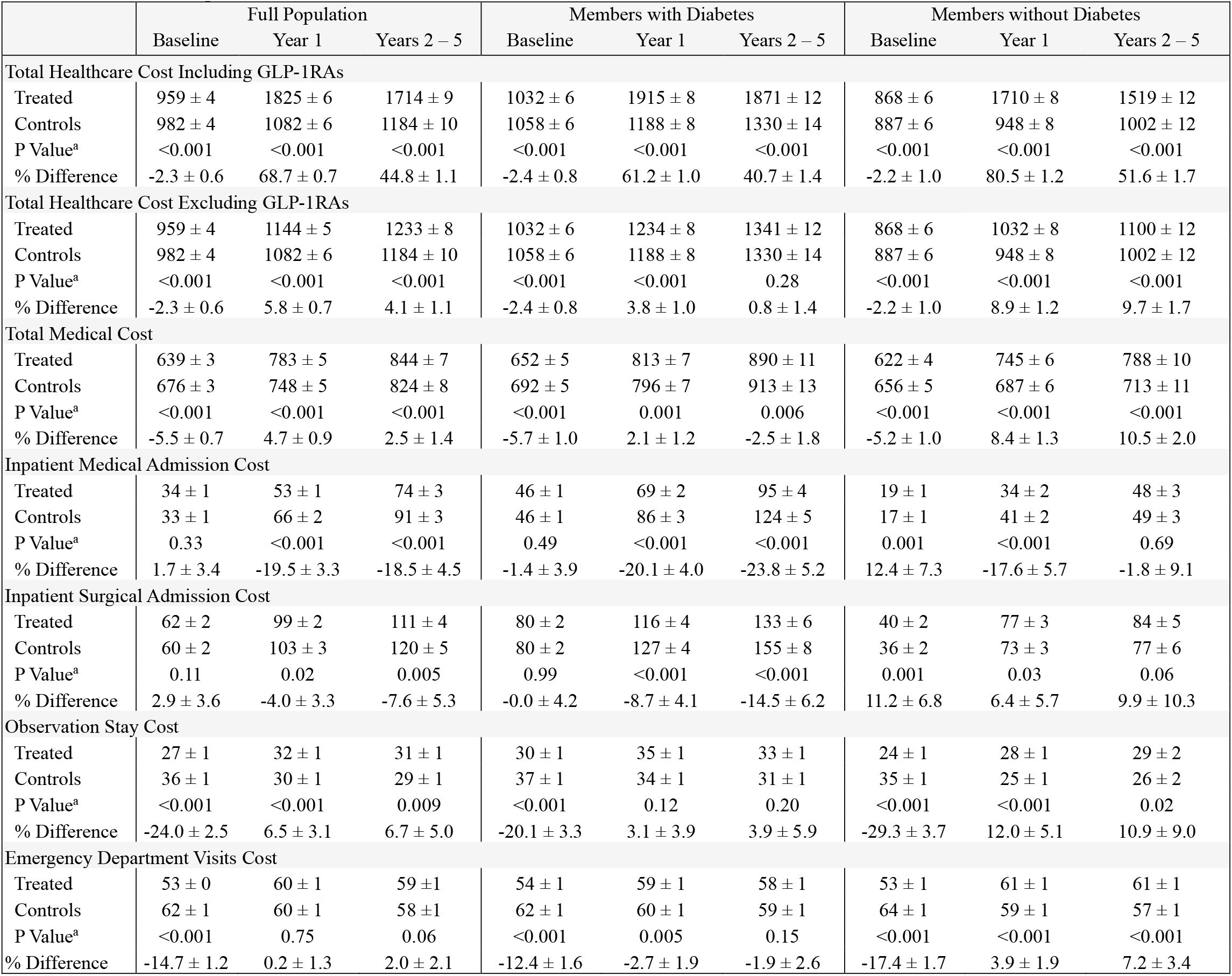
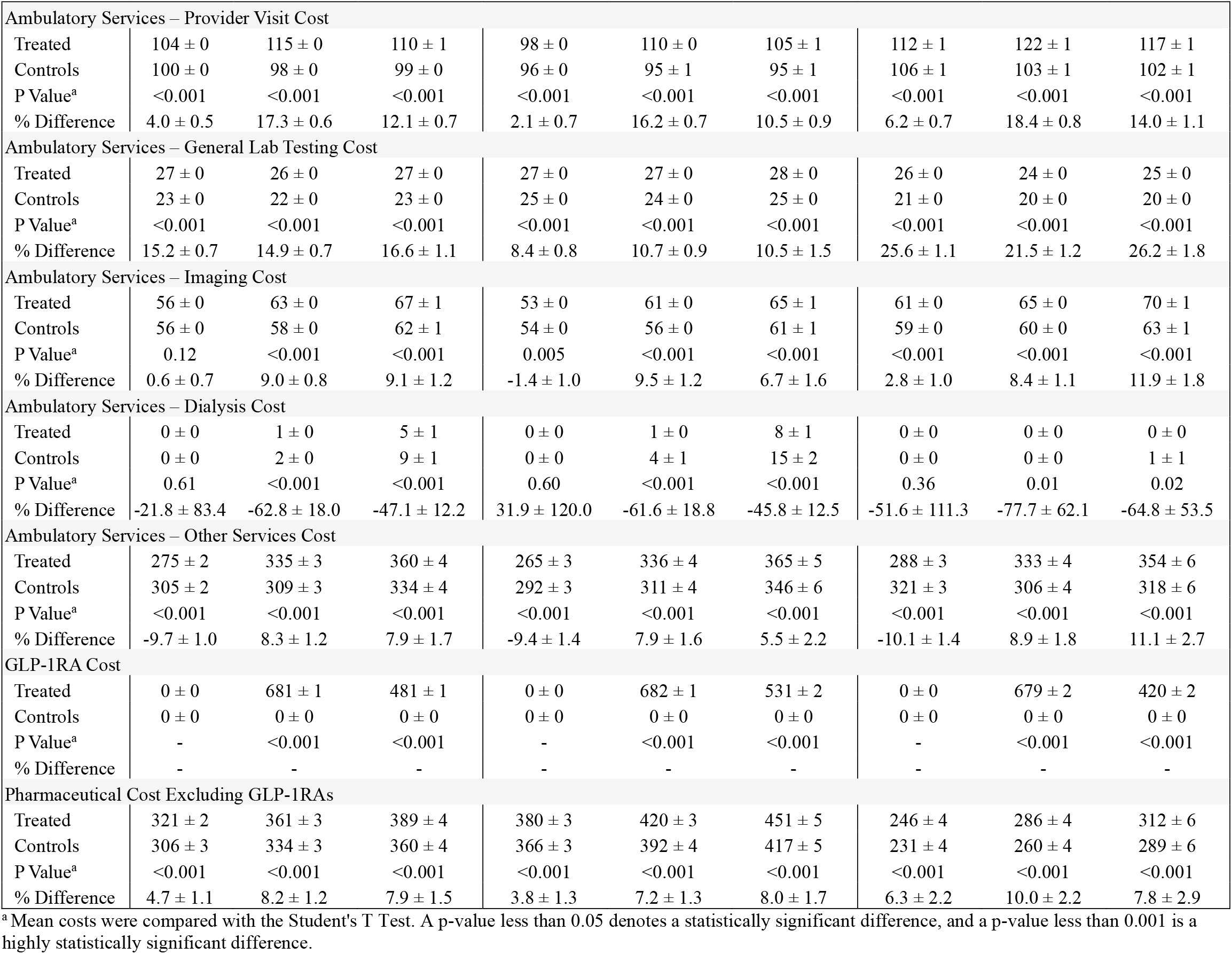
Baseline and Follow-up Healthcare Costs ($ PMPM ± 95% CI) and Cost Differences (% ± 95% CI).

|  | Full Population |  |  | Members with Diabetes |  |  | Members without Diabetes |  |  |
| --- | --- | --- | --- | --- | --- | --- | --- | --- | --- |
|  | Baseline | Year 1 | Years 2 – 5 | Baseline | Year 1 | Years 2 – 5 | Baseline | Year 1 | Years 2 – 5 |
| **Total Healthcare Cost Including GLP-1RAs** |  |  |  |  |  |  |  |  |  |
| Treated | 959 ± 4 | 1825 ± 6 | 1714 ± 9 | 1032 ± 6 | 1915 ± 8 | 1871 ± 12 | 868 ± 6 | 1710 ± 8 | 1519 ± 12 |
| Controls | 982 ± 4 | 1082 ± 6 | 1184 ± 10 | 1058 ± 6 | 1188 ± 8 | 1330 ± 14 | 887 ± 6 | 948 ± 8 | 1002 ± 12 |
| P Value<sup>a</sup> | <0.001 | <0.001 | <0.001 | <0.001 | <0.001 | <0.001 | <0.001 | <0.001 | <0.001 |
| % Difference | -2.3 ± 0.6 | 68.7 ± 0.7 | 44.8 ± 1.1 | -2.4 ± 0.8 | 61.2 ± 1.0 | 40.7 ± 1.4 | -2.2 ± 1.0 | 80.5 ± 1.2 | 51.6 ± 1.7 |
| **Total Healthcare Cost Excluding GLP-1RAs** |  |  |  |  |  |  |  |  |  |
| Treated | 959 ± 4 | 1144 ± 5 | 1233 ± 8 | 1032 ± 6 | 1234 ± 8 | 1341 ± 12 | 868 ± 6 | 1032 ± 8 | 1100 ± 12 |
| Controls | 982 ± 4 | 1082 ± 6 | 1184 ± 10 | 1058 ± 6 | 1188 ± 8 | 1330 ± 14 | 887 ± 6 | 948 ± 8 | 1002 ± 12 |
| P Value<sup>a</sup> | <0.001 | <0.001 | <0.001 | <0.001 | <0.001 | 0.28 | <0.001 | <0.001 | <0.001 |
| % Difference | -2.3 ± 0.6 | 5.8 ± 0.7 | 4.1 ± 1.1 | -2.4 ± 0.8 | 3.8 ± 1.0 | 0.8 ± 1.4 | -2.2 ± 1.0 | 8.9 ± 1.2 | 9.7 ± 1.7 |
| **Total Medical Cost** |  |  |  |  |  |  |  |  |  |
| Treated | 639 ± 3 | 783 ± 5 | 844 ± 7 | 652 ± 5 | 813 ± 7 | 890 ± 11 | 622 ± 4 | 745 ± 6 | 788 ± 10 |
| Controls | 676 ± 3 | 748 ± 5 | 824 ± 8 | 692 ± 5 | 796 ± 7 | 913 ± 13 | 656 ± 5 | 687 ± 6 | 713 ± 11 |
| P Value<sup>a</sup> | <0.001 | <0.001 | <0.001 | <0.001 | 0.001 | 0.006 | <0.001 | <0.001 | <0.001 |
| % Difference | -5.5 ± 0.7 | 4.7 ± 0.9 | 2.5 ± 1.4 | -5.7 ± 1.0 | 2.1 ± 1.2 | -2.5 ± 1.8 | -5.2 ± 1.0 | 8.4 ± 1.3 | 10.5 ± 2.0 |
| **Inpatient Medical Admission Cost** |  |  |  |  |  |  |  |  |  |
| Treated | 34 ± 1 | 53 ± 1 | 74 ± 3 | 46 ± 1 | 69 ± 2 | 95 ± 4 | 19 ± 1 | 34 ± 2 | 48 ± 3 |
| Controls | 33 ± 1 | 66 ± 2 | 91 ± 3 | 46 ± 1 | 86 ± 3 | 124 ± 5 | 17 ± 1 | 41 ± 2 | 49 ± 3 |
| P Value<sup>a</sup> | 0.33 | <0.001 | <0.001 | 0.49 | <0.001 | <0.001 | 0.001 | <0.001 | 0.69 |
| % Difference | 1.7 ± 3.4 | -19.5 ± 3.3 | -18.5 ± 4.5 | -1.4 ± 3.9 | -20.1 ± 4.0 | -23.8 ± 5.2 | 12.4 ± 7.3 | -17.6 ± 5.7 | -1.8 ± 9.1 |
| **Inpatient Surgical Admission Cost** |  |  |  |  |  |  |  |  |  |
| Treated | 62 ± 2 | 99 ± 2 | 111 ± 4 | 80 ± 2 | 116 ± 4 | 133 ± 6 | 40 ± 2 | 77 ± 3 | 84 ± 5 |
| Controls | 60 ± 2 | 103 ± 3 | 120 ± 5 | 80 ± 2 | 127 ± 4 | 155 ± 8 | 36 ± 2 | 73 ± 3 | 77 ± 6 |
| P Value<sup>a</sup> | 0.11 | 0.02 | 0.005 | 0.99 | <0.001 | <0.001 | 0.001 | 0.03 | 0.06 |
| % Difference | 2.9 ± 3.6 | -4.0 ± 3.3 | -7.6 ± 5.3 | -0.0 ± 4.2 | -8.7 ± 4.1 | -14.5 ± 6.2 | 11.2 ± 6.8 | 6.4 ± 5.7 | 9.9 ± 10.3 |
| **Observation Stay Cost** |  |  |  |  |  |  |  |  |  |
| Treated | 27 ± 1 | 32 ± 1 | 31 ± 1 | 30 ± 1 | 35 ± 1 | 33 ± 1 | 24 ± 1 | 28 ± 1 | 29 ± 2 |
| Controls | 36 ± 1 | 30 ± 1 | 29 ± 1 | 37 ± 1 | 34 ± 1 | 31 ± 1 | 35 ± 1 | 25 ± 1 | 26 ± 2 |
| P Value<sup>a</sup> | <0.001 | <0.001 | 0.009 | <0.001 | 0.12 | 0.20 | <0.001 | <0.001 | 0.02 |
| % Difference | -24.0 ± 2.5 | 6.5 ± 3.1 | 6.7 ± 5.0 | -20.1 ± 3.3 | 3.1 ± 3.9 | 3.9 ± 5.9 | -29.3 ± 3.7 | 12.0 ± 5.1 | 10.9 ± 9.0 |
| **Emergency Department Visits Cost** |  |  |  |  |  |  |  |  |  |
| Treated | 53 ± 0 | 60 ± 1 | 59 ± 1 | 54 ± 1 | 59 ± 1 | 58 ± 1 | 53 ± 1 | 61 ± 1 | 61 ± 1 |
| Controls | 62 ± 1 | 60 ± 1 | 58 ± 1 | 62 ± 1 | 60 ± 1 | 59 ± 1 | 64 ± 1 | 59 ± 1 | 57 ± 1 |
| P Value<sup>a</sup> | <0.001 | 0.75 | 0.06 | <0.001 | 0.005 | 0.15 | <0.001 | <0.001 | <0.001 |
| % Difference | -14.7 ± 1.2 | 0.2 ± 1.3 | 2.0 ± 2.1 | -12.4 ± 1.6 | -2.7 ± 1.9 | -1.9 ± 2.6 | -17.4 ± 1.7 | 3.9 ± 1.9 | 7.2 ± 3.4 |

**Table 2. Continued**
|  | Full Population |  |  | Members with Diabetes |  |  | Members without Diabetes |  |  |
| --- | --- | --- | --- | --- | --- | --- | --- | --- | --- |
|  | Baseline | Year 1 | Years 2 – 5 | Baseline | Year 1 | Years 2 – 5 | Baseline | Year 1 | Years 2 – 5 |
| **Ambulatory Services – Provider Visit Cost** |  |  |  |  |  |  |  |  |  |
| Treated | 104 ± 0 | 115 ± 0 | 110 ± 1 | 98 ± 0 | 110 ± 0 | 105 ± 1 | 112 ± 1 | 122 ± 1 | 117 ± 1 |
| Controls | 100 ± 0 | 98 ± 0 | 99 ± 0 | 96 ± 0 | 95 ± 1 | 95 ± 1 | 106 ± 1 | 103 ± 1 | 102 ± 1 |
| P Value<sup>a</sup> | <0.001 | <0.001 | <0.001 | <0.001 | <0.001 | <0.001 | <0.001 | <0.001 | <0.001 |
| % Difference | 4.0 ± 0.5 | 17.3 ± 0.6 | 12.1 ± 0.7 | 2.1 ± 0.7 | 16.2 ± 0.7 | 10.5 ± 0.9 | 6.2 ± 0.7 | 18.4 ± 0.8 | 14.0 ± 1.1 |
| **Ambulatory Services – General Lab Testing Cost** |  |  |  |  |  |  |  |  |  |
| Treated | 27 ± 0 | 26 ± 0 | 27 ± 0 | 27 ± 0 | 27 ± 0 | 28 ± 0 | 26 ± 0 | 24 ± 0 | 25 ± 0 |
| Controls | 23 ± 0 | 22 ± 0 | 23 ± 0 | 25 ± 0 | 24 ± 0 | 25 ± 0 | 21 ± 0 | 20 ± 0 | 20 ± 0 |
| P Value<sup>a</sup> | <0.001 | <0.001 | <0.001 | <0.001 | <0.001 | <0.001 | <0.001 | <0.001 | <0.001 |
| % Difference | 15.2 ± 0.7 | 14.9 ± 0.7 | 16.6 ± 1.1 | 8.4 ± 0.8 | 10.7 ± 0.9 | 10.5 ± 1.5 | 25.6 ± 1.1 | 21.5 ± 1.2 | 26.2 ± 1.8 |
| **Ambulatory Services – Imaging Cost** |  |  |  |  |  |  |  |  |  |
| Treated | 56 ± 0 | 63 ± 0 | 67 ± 1 | 53 ± 0 | 61 ± 0 | 65 ± 1 | 61 ± 0 | 65 ± 0 | 70 ± 1 |
| Controls | 56 ± 0 | 58 ± 0 | 62 ± 1 | 54 ± 0 | 56 ± 0 | 61 ± 1 | 59 ± 0 | 60 ± 0 | 63 ± 1 |
| P Value<sup>a</sup> | 0.12 | <0.001 | <0.001 | 0.005 | <0.001 | <0.001 | <0.001 | <0.001 | <0.001 |
| % Difference | 0.6 ± 0.7 | 9.0 ± 0.8 | 9.1 ± 1.2 | -1.4 ± 1.0 | 9.5 ± 1.2 | 6.7 ± 1.6 | 2.8 ± 1.0 | 8.4 ± 1.1 | 11.9 ± 1.8 |
| **Ambulatory Services – Dialysis Cost** |  |  |  |  |  |  |  |  |  |
| Treated | 0 ± 0 | 1 ± 0 | 5 ± 1 | 0 ± 0 | 1 ± 0 | 8 ± 1 | 0 ± 0 | 0 ± 0 | 0 ± 0 |
| Controls | 0 ± 0 | 2 ± 0 | 9 ± 1 | 0 ± 0 | 4 ± 1 | 15 ± 2 | 0 ± 0 | 0 ± 0 | 1 ± 1 |
| P Value<sup>a</sup> | 0.61 | <0.001 | <0.001 | 0.60 | <0.001 | <0.001 | 0.36 | 0.01 | 0.02 |
| % Difference | -21.8 ± 83.4 | -62.8 ± 18.0 | -47.1 ± 12.2 | 31.9 ± 120.0 | -61.6 ± 18.8 | -45.8 ± 12.5 | -51.6 ± 111.3 | -77.7 ± 62.1 | -64.8 ± 53.5 |
| **Ambulatory Services – Other Services Cost** |  |  |  |  |  |  |  |  |  |
| Treated | 275 ± 2 | 335 ± 3 | 360 ± 4 | 265 ± 3 | 336 ± 4 | 365 ± 5 | 288 ± 3 | 333 ± 4 | 354 ± 6 |
| Controls | 305 ± 2 | 309 ± 3 | 334 ± 4 | 292 ± 3 | 311 ± 4 | 346 ± 6 | 321 ± 3 | 306 ± 4 | 318 ± 6 |
| P Value<sup>a</sup> | <0.001 | <0.001 | <0.001 | <0.001 | <0.001 | <0.001 | <0.001 | <0.001 | <0.001 |
| % Difference | -9.7 ± 1.0 | 8.3 ± 1.2 | 7.9 ± 1.7 | -9.4 ± 1.4 | 7.9 ± 1.6 | 5.5 ± 2.2 | -10.1 ± 1.4 | 8.9 ± 1.8 | 11.1 ± 2.7 |
| **GLP-1RA Cost** |  |  |  |  |  |  |  |  |  |
| Treated | 0 ± 0 | 681 ± 1 | 481 ± 1 | 0 ± 0 | 682 ± 1 | 531 ± 2 | 0 ± 0 | 679 ± 2 | 420 ± 2 |
| Controls | 0 ± 0 | 0 ± 0 | 0 ± 0 | 0 ± 0 | 0 ± 0 | 0 ± 0 | 0 ± 0 | 0 ± 0 | 0 ± 0 |
| P Value<sup>a</sup> | - | <0.001 | <0.001 | - | <0.001 | <0.001 | - | <0.001 | <0.001 |
| % Difference | - | - | - | - | - | - | - | - | - |
| **Pharmaceutical Cost Excluding GLP-1RAs** |  |  |  |  |  |  |  |  |  |
| Treated | 321 ± 2 | 361 ± 3 | 389 ± 4 | 380 ± 3 | 420 ± 3 | 451 ± 5 | 246 ± 4 | 286 ± 4 | 312 ± 6 |
| Controls | 306 ± 3 | 334 ± 3 | 360 ± 4 | 366 ± 3 | 392 ± 4 | 417 ± 5 | 231 ± 4 | 260 ± 4 | 289 ± 6 |
| P Value<sup>a</sup> | <0.001 | <0.001 | <0.001 | <0.001 | <0.001 | <0.001 | <0.001 | <0.001 | <0.001 |
| % Difference | 4.7 ± 1.1 | 8.2 ± 1.2 | 7.9 ± 1.5 | 3.8 ± 1.3 | 7.2 ± 1.3 | 8.0 ± 1.7 | 6.3 ± 2.2 | 10.0 ± 2.2 | 7.8 ± 2.9 |

<sup>a</sup> Mean costs were compared with the Student's T Test. A p-value less than 0.05 denotes a statistically significant difference, and a p-value less than 0.001 is a highly statistically significant difference.

**Fig 1.**
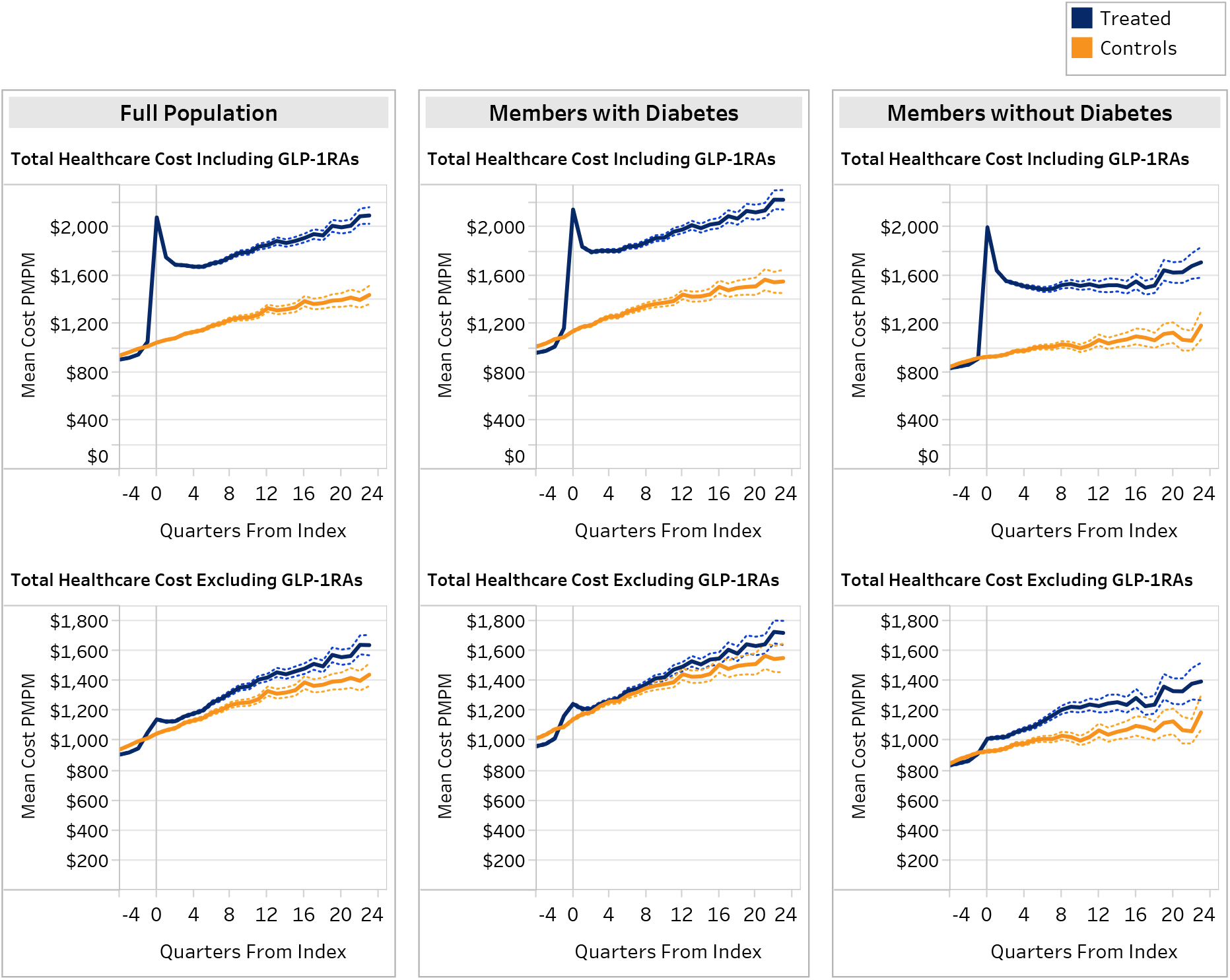
Healthcare Costs* in Quarter Relative to Initial Prescribing of GLP-1RA: Treated and Control Groups, and for Members with and without Diabetes. **\***Costs include both facility and professional claim types. Dotted lines are 95% confidence intervals.

**Fig. 2.**
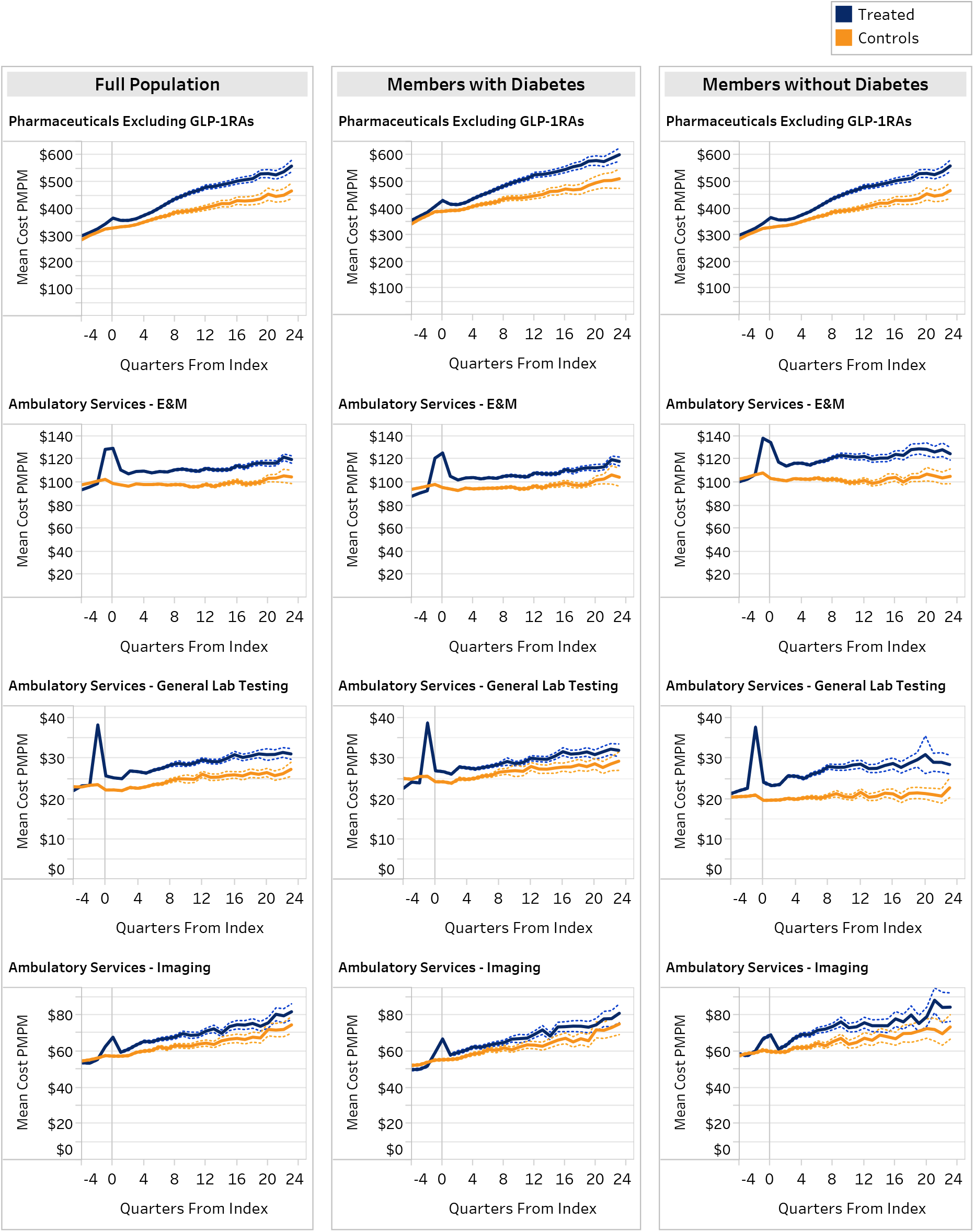
Select Cost and Utilization Categories* for GLP-1RA Treated and Control Groups, Relative to Quarter of Initial Prescribing of GLP-1RA. *Costs include both facility and professional claim types. Dotted lines are 95% confidence intervals.

**Fig 3.**
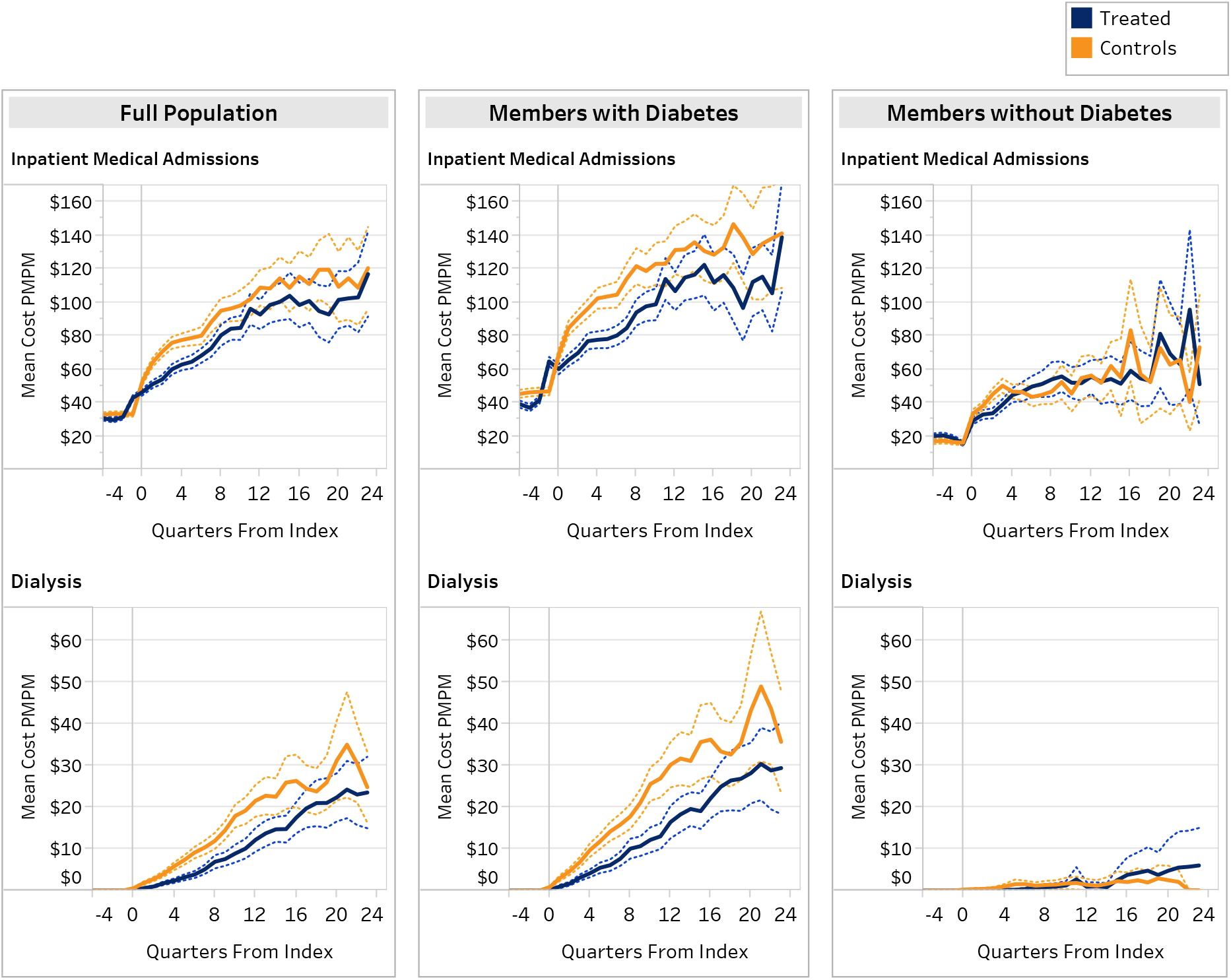
Inpatient Medical Admissions and Dialysis Costs* for GLP-1RA Treated and Control Groups, Relative to Quarter of Initial Prescribing of GLP-1RA. **\***Costs include both facility and professional claim types. Dotted lines are 95% confidence intervals.

Immediately prior to and after GLP-1RA initiation, total costs spiked for all treated members and for those with and without diabetes (Figures 1-3). The rise prior to GLP-1RA initiation was primarily in ambulatory provider visits and general laboratory testing services. After initiation, the largest component of the cost increases in the treated groups was the GLP-1RAs themselves.

In follow-up year 1, excluding costs of the GLP-1RA, there was a 5.8% (95% CI: 5.0–6.5%; p<0.001) increase in medical and non-GLP-1RA pharmacy costs for the total treated population and 4.1% (95% CI: 3.1–5.2%; p<0.001) in years 2-6 (Table 2), increases largely driven by costs in members without diabetes (Table 3). Treated members with diabetes had 3.8% (95% CI: 2.8–4.8%; p<0.001) higher medical and non-GLP-1RA pharmacy costs in the first year of follow-up and 0.8% higher in years 2-6 (95% CI: -0.6–2.1%; p=0.278). There were several categories of care where treated members with diabetes had significantly higher non-GLP-1RA costs: ambulatory provider visits (16.2% in year 1 of follow-up, 10.5% in years 2-6), general laboratory services (10.7% in the first year of follow-up and 10.5% in years 2-6), and imaging (9.5% in the first year of follow-up and 6.7% in years 2-6). These were largely offset by 20.1–23.8% lower inpatient costs and 45.8–61.6% lower dialysis costs (both p<0.001).

Treated members without diabetes medical and non-GLP-1RA pharmacy costs were 8.9% (95% CI: 7.7%–10.0%; p<0.001) higher in the first year of follow-up, rising to 9.7% (95% CI: 8.0%–11.5%; p<0.001) in years 2-6. Overall medical costs (non-pharmacy) were 8.4-10.5% higher in follow up years (p<0.001). Higher costs for treated members without diabetes were seen in all medical categories except inpatient medical DRGs, where there were significantly lower costs in the first follow-up year but not in years 2-6. The largest increases in costs were found in ambulatory services including provider visits, imaging and laboratory services.

Treatment with newer generation GLP1-RAs (semaglutide and tirzepatide) was more common than treatment with older generation agents, and associated healthcare costs were substantially lower (Table S4 and Figures S1-S3).

Excluding costs of the GLP-1RAs themselves, medical and pharmacy costs for members with diabetes treated with newer generation GLP-1RAs were 4.8% lower in years 2-6 than controls, while they were 10% higher for older GLP-1RAs. In members without diabetes, costs for the newer generation GLP-1RAs were 6.8-7.2% higher, while those for older GLP-1RAs were 15.6-20% higher (Table S5). For members without diabetes, costs associated with gastrointestinal conditions, the most common side effect of GLP-1RAs, were substantially higher than in controls for both newer and older generation of GLP-1RAs (Table S6).

## Discussion

Our study is one of the largest published on the real-world costs associated with GLP-1RAs. We find that increases in health care costs associated with treatment extend beyond the cost of the agents themselves, with differing effects for members with diabetes and those without. Total costs including GLP-1RAs were substantially higher for all treated members. Once excluding the costs of the GLP-1RA agents, members with diabetes had a modest increase in costs in the first year of treatment only. The increased costs for non-GLP-1RA drugs and common outpatient services associated with treating members with diabetes were offset by large decreases in costs for inpatient medical admissions and dialysis. For members treated with newer generation GLP-1RAs, the overall costs of medical care were lower once excluding the costs of the GLP-1RA medications. Our findings for treated members without diabetes were very different. Treatment with GLP-1RA for members without diabetes was associated with a 9-10% increase in costs during the six-year follow-up. Treated members without diabetes had higher costs in almost every category of care. While the associated costs were much larger with older generation GLP1-RAs, we still found that costs for members without diabetes treated with newer agents were 6-7% higher than control members, excluding the costs of the GLP-1RAs.

Prior studies of the impact of GLP-1RAs on the cost of medical care have primarily been actuarial studies or are cost-effectiveness models. The actuarial studies have shown that GLP-1RAs have been responsible for a large proportion of the recent increases in expenditures at the plan and employer level.^13,20^ The cost-effectiveness studies have found different impacts of GLP-1RAs based on indication for treatment. For people with diabetes GLP-1RAs were either cost-saving or cost-effective depending upon the agent using a threshold of $100,000/quality adjusted life year (QALY).^25^ For people with obesity, GLP-1RAs have not been found to be cost-effective, with studies suggesting that discounts of up to 82% of current pricing would be required to approach a $100,000/QALY threshold.^16,26^ Importantly, these studies suggested either minimal increases in non-GLP-1RA medical costs, or a substantial reduction in these costs. A more recent study used electronic medical record and found that treatment with GLP-1RAs was associated with increased costs beyond the costs of the GLP1-RAs themselves, however, this study did not have a control, used Medicare reference pricing for costs and only measured services rendered within the health care system.^21^

Several limitations of our study should be considered. Our study uses carefully constructed matching to identify our control population; however, it is still a quasi-experimental design. The findings that most of the increased costs in people without diabetes was found in services associated with increased contacts with the health care systems, that newer generation GLP-1RAs with lower side-effect profiles were associated with lower costs, and that a substantial proportion of the increased costs are for claims with these side effects lends credence to the causal association of GLP-1RA treatment and increased costs. Our follow-up time was 6 years. While there could be additional health improvements in longer follow-up, given the average turnover in commercial health plan populations, such improvements, if found, would accrue to other parties. While the size of treated population without diabetes with 4-6 follow-up diminished, it remained large enough to result in stable measurement for most categories of care (Table S7).We did not have access to data on pharmacy rebates or other incentives in the drug management industry. While these could reduce the total costs of GLP-1RA coverage, they would not affect the costs of other medical services. Our data did not allow us to measure the impact of GLP-1RAs on quality of life, an important outcome of treatment, particularly for those without diabetes. Finally, GLP-1RAs can be obtained outside of insurance potentially leading to misclassification of untreated populations. Such misclassification would result in higher costs in the untreated and therefore diminish the differences between treated and controls.

Our large real-world study of GLP-1RAs in commercially insured adults has two primary health policy implications. Our study supports the use of GLP-1RAs for treating people with diabetes. Beyond the agents themselves, the modest increases in costs in the initial year of treatment and cost-neutrality in years 2-6 due to large decreases in inpatient admissions and dialysis costs are notable. If confirmed elsewhere, this would support the broader use of these agents in treating diabetes from both health and health care cost perspectives. For people without diabetes, most of whom are likely to have obesity, our study suggests that even if GLP-1RAs agents were available for free, there would still be a significant increase in total health care costs. With an estimated 40% of the US adult population eligible for obesity treatment with GLP-1RAs, the costs of covering treatment for this indication would have a substantial impact on insurance premiums.^27,28^

## Data Availability

Individual participant data will not be made available, in compliance with the data use agreement with the Blue Cross Blue Shield Association.

## Author Contributions

AM and CM had full access to all of the data in the study and take responsibility for the integrity of the data and the accuracy of the data analysis.

## Concept and design

DW, HC, AM, CM

## Acquisition, analysis, or interpretation of data

DW, AM, CM, HC, JS

## Drafting of the manuscript

DW, AM, CM, LAT

## Critical review of the manuscript for important intellectual content

HC, RH, JS, VV, SH, SR, MM, JS, LAT

## Statistical analysis

AM, CM

## Obtained funding

HC

## Administrative, technical, or material support

RH, JK, VV, SH, LAT

## Supervision

DW, HC

## Disclaimer

The content is solely the responsibility of the authors and does not necessarily represent the official views of the funding entity.

## Additional Contributions

The authors acknowledge Bob Darin (concept, design, review), Stephanie Tomlin (review, editing), and Thomas Parakilatu, Neha Jajodia, and Jen Zhong (data management) for their valuable contributions to this project.

