## Supplementary Tables and Figures for "The Real-World Costs of GLP-1 Receptor Agonist Treatment"

| **Table of Contents** |
|---|
| Table S1a: GLP-1RA user cohort identification and retention |
| Table S1b: non-GLP-1RA user cohort identification and retention |
| Table S2: Pre-matching characteristics of treated and control cohorts |
| Table S3: Baseline and follow-up all-cause healthcare utilization in matched GLP-1RA treated and control cohorts |
| Table S4: Post-matching population sizes by GLP-1RA treatment |
| Table S5: Baseline and follow-up all-cause healthcare costs in matched treated and control cohorts by generation of GLP-1RAs |
| Table S6: Follow-up gastrointestinal side effect-related medical and pharmacy costs in matched treated and control cohorts, by index GLP-1RA type |
| Table S7: Follow-up time of matched treated and control cohorts by quarter |
| Figure S1. Healthcare costs in quarter relative to initial prescribing of GLP-1RA: by GLP-1RA generation and diabetes status |
| Figure S2. Costs for select utilization categories in quarter relative to initial prescribing of GLP-1RA: by GLP-1RA generation and diabetes status |
| Figure S3. Inpatient medical admissions and dialysis costs in quarter relative to initial prescribing of GLP-1RA: by GLP-1RA generation and diabetes status |

| Table S1a: treated cohort identification and retention | | |
|---|---|---|
| Criteria | Patients remaining | |
| | N | % of starting population |
| Individuals aged 18-64 years (with nonmissing gender, date of birth, and zip code) who had their first pharmacy claim for any GLP1 product (tirzepatide, albiglutide, semaglutide, liraglutide, lixisenatide, dulaglutide, or exenatide) between January 1st, 2017, and February 29th, 2024. The date of this claim is the index date and individuals must have primary medical and pharmacy coverage. | 1,725,097 | 100.0% |
| Minimum of 12 months of continuous enrollment in primary medical and pharmacy coverage prior to index date and 6 months of continuous enrollment in primary medical and pharmacy coverage on and following index date | 1,018,206 | 59.0% |
| Individuals remaining after exclusions (individuals having any of the following conditions within 1 year prior to index: cancer, transplant, hypoglycemia with hospitalization, dementia, ESRD diagnosis or dialysis use (home or facility based) , respiratory failure, end-stage liver disease, hospice stay, nursing home stay, mechanical ventilation, psychosis, anorexia, bulimia, pregnancy, bariatric surgery) | 936,777 | 54.3% |

| Table S1b: non-treated cohort identification and retention | | |
|---|---|---|
| Criteria | Patients remaining | |
| | N | % of starting population |
| Individuals aged 18-64 years of age (with nonmissing gender, date of birth, and zip code) who had no evidence of exposure to any GLP-1RA product (tirzepatide, albiglutide, semaglutide, liraglutide, lixisenatide, dulaglutide, or exenatide) between January 1st, 2015, and August 31st, 2024. The pseudo-index date for each non-user is randomly assigned based on the distribution of feasible index dates among members in the same age bracket who had primary medical and pharmacy coverage in the same quarter. | 23,261,958 | 100.0% |
| Minimum of 12 months of continuous enrollment in primary medical and pharmacy coverage prior to pseudo-index date and 6 months of continuous enrollment in primary medical and pharmacy coverage on and following pseudo-index date | 22,716,175 | 97.7% |
| Individuals remaining after exclusions (individuals having any of the following conditions within 1 year prior to index: cancer, transplant, hypoglycemia with hospitalization, dementia, ESRD diagnosis or dialysis use (home or facility based) , respiratory failure, end-stage liver disease, hospice stay, nursing home stay, mechanical ventilation, psychosis, anorexia, bulimia, pregnancy, bariatric surgery) | 18,281,409 | 78.6% |

**Table S2: Pre-matching characteristics of treated and control cohorts**

| Characteristic considered in the matching algorithm | Full Population (Pre-Matching) | | |
|---|---|---|---|
| | Treated | Controls | SMD |
| **Age at Index (Years), mean (SD)** | 32.8 (9.7) | 24.8 (13.7) | 0.67 |
| **Gender (% Female)** | 60.4% | 52.1% | 0.17 |
| **Race/Ethnicity** | | | |
| White | 66.0% | 68.3% | 0.15 |
| Black | 8.1% | 5.5% | |
| Hispanic | 9.3% | 7.6% | |
| Other | 3.7% | 5.8% | |
| Unknown | 12.9% | 12.8% | |
| Type 1 Diabetes | 1.6% | 0.5% | 0.11 |
| Type 2 Diabetes with complications | 27.6% | 1.2% | 0.81 |
| Type 2 Diabetes without complications | 35.3% | 3.0% | 0.90 |
| Obesity | 44.7% | 9.1% | 0.89 |
| Liver Disease | 0.8% | 0.4% | 0.06 |
| Hypertension | 54.8% | 15.5% | 0.90 |
| Anxiety | 15.8% | 10.0% | 0.17 |
| Depression | 12.2% | 6.4% | 0.20 |
| Heart Failure | 1.5% | 0.3% | 0.13 |
| Coronary Artery Disease | 5.3% | 1.2% | 0.23 |
| Chronic Kidney Disease | 3.2% | 0.5% | 0.20 |
| Obstructive Sleep Apnea | 11.9% | 2.3% | 0.38 |
| Prior Treatment | | | |
| Insulin | 12.0% | 0.8% | 0.47 |
| SGLT2 Inhibitors | 14.8% | 0.5% | 0.56 |
| Sulfonylureas[a] | 17.0% | 0.7% | 0.60 |
| DPP-4 Inhibitors | 9.6% | 0.3% | 0.44 |
| Biguanides | 48.2% | 3.0% | 1.21 |
| Number of Non-Bariatric Inpatient Stays | | | |
| <0.001 | 95.6% | 98.1% | 0.14 |
| 1 | 3.6% | 1.6% | |
| 2+ | 0.7% | 0.3% | |
| Number of other prescription classes used at baseline | | | |
| <0.001 | 4.4% | 18.4% | 0.77 |
| 1 to 3 | 25.9% | 43.3% | |
| 4 to 6 | 28.3% | 22.5% | |
| 7 to 9 | 19.6% | 9.5% | |
| 10+ | 21.8% | 6.3% | |

a The sulfonylureas category also includes meglitinide analogues and thiazolidinediones.

**Table S3: Baseline and follow-up all-cause healthcare utilization in matched GLP-1RA treated and control cohortsa**

| Mean (95% confidence interval) of encounters per 1,000 member months[b] | Full Population | | | Members with Diabetes | | | Members without Diabetes | | |
|---|---|---|---|---|---|---|---|---|---|
| | Baseline | Year 1 | Years 2-6 | Baseline | Year 1 | Years 2-6 | Baseline | Year 1 | Years 2-6 |
| ***Inpatient medical admissions*** [c] | | | | | | | | | |
| *Treated* | 2.5 ± 0.0 | 3.2 ± 0.1 | 4.0 ± 0.1 | 3.3 ± 0.1 | 4.1 ± 0.1 | 5.0 ± 0.1 | 1.4 ± 0.1 | 2.2 ± 0.1 | 2.9 ± 0.2 |
| *Controls* | 2.4 ± 0.0 | 4.0 ± 0.1 | 4.9 ± 0.1 | 3.3 ± 0.1 | 5.0 ± 0.1 | 6.4 ± 0.2 | 1.3 ± 0.1 | 2.7 ± 0.1 | 3.1 ± 0.1 |
| *P-value* | 0.002 | <0.001 | <0.001 | 0.208 | <0.001 | <0.001 | 0.000 | <0.001 | 0.023 |
| *% difference [95% CI]* | 4.1% ± 2.7% | -19.2% ± 2.2% | -18.1% ± 3.2% | 1.8% ± 2.9% | -18.9% ± 2.6% | -22.0% ± 3.5% | 11.5% ± 6.3% | -19.8% ± 4.1% | -8.1% ± 7.0% |
| ***Inpatient surgical admissions*** [c] | | | | | | | | | |
| *Treated* | 1.6 ± 0.0 | 2.3 ± 0.0 | 2.5 ± 0.1 | 1.6 ± 0.0 | 2.6 ± 0.1 | 2.8 ± 0.1 | 1.6 ± 0.0 | 1.9 ± 0.0 | 2.2 ± 0.1 |
| *Controls* | 1.5 ± 0.0 | 2.4 ± 0.0 | 2.5 ± 0.1 | 1.5 ± 0.0 | 2.8 ± 0.1 | 3.0 ± 0.1 | 1.5 ± 0.0 | 1.9 ± 0.0 | 1.9 ± 0.1 |
| *P-value* | 0.102 | <0.001 | 0.312 | 0.102 | <0.001 | 0.016 | 0.102 | 0.412 | <0.001 |
| *% difference [95% CI]* | 2.1% ± 2.6% | -3.7% ± 2.2% | 1.9% ± 3.6% | 2.1% ± 2.6% | -6.5% ± 2.7% | -5.5% ± 4.4% | 2.1% ± 2.6% | 1.5% ± 3.6% | 16.4% ± 6.3% |
| ***Emergency department visits*** | | | | | | | | | |
| *Treated* | 22.7 ± 0.1 | 24.0 ± 0.1 | 23.7 ± 0.2 | 23.3 ± 0.2 | 24.3 ± 0.2 | 24.0 ± 0.3 | 21.9 ± 0.2 | 23.6 ± 0.2 | 23.4 ± 0.3 |
| *Controls* | 26.2 ± 0.2 | 24.9 ± 0.2 | 24.1 ± 0.2 | 26.1 ± 0.2 | 25.6 ± 0.2 | 25.3 ± 0.3 | 26.2 ± 0.2 | 24.1 ± 0.2 | 22.7 ± 0.3 |
| *P-value* | <0.001 | <0.001 | 0.015 | <0.001 | <0.001 | <0.001 | <0.001 | 0.005 | 0.008 |
| *% difference [95% CI]* | -13.4% ± 0.8% | -3.8% ± 0.9% | -1.6% ± 1.3% | -10.9% ± 1.1% | -5.2% ± 1.2% | -4.9% ± 1.7% | -16.6% ± 1.2% | -1.9% ± 1.3% | 2.9% ± 2.1% |
| ***Ambulatory provider visits*** | | | | | | | | | |
| *Treated* | 754.8 ± 1.7 | 842.2 ± 1.8 | 795.2 ± 2.2 | 716.9 ± 2.1 | 807.4 ± 2.3 | 762.3 ± 2.8 | 802.5 ± 2.8 | 885.9 ± 3.0 | 835.9 ± 3.6 |
| *Controls* | 737.0 ± 1.7 | 716.1 ± 1.8 | 705.6 ± 2.3 | 701.3 ± 2.1 | 687.4 ± 2.3 | 678.7 ± 2.9 | 781.8 ± 2.8 | 752.0 ± 2.9 | 738.8 ± 3.7 |
| *P-value* | <0.001 | <0.001 | <0.001 | <0.001 | <0.001 | <0.001 | <0.001 | <0.001 | <0.001 |
| *% difference [95% CI]* | 2.4% ± 0.3% | 17.6% ± 0.4% | 12.7% ± 0.5% | 2.2% ± 0.4% | 17.5% ± 0.5% | 12.3% ± 0.6% | 2.7% ± 0.5% | 17.8% ± 0.6% | 13.1% ± 0.7% |

[a] Members included in this table were required to have at least 12 months of baseline and at least 6 months of follow-up continuous enrollment. Utilization is assigned to the time period based on the start date of the stay, procedure, or prescription fill.

[b] Mean encounters per 1,000 member months were compared with the Student's T Test. A p-value less than 0.05 denotes a statistically significant difference, and a p-value less than 0.001 is a highly statistically significant difference.

[c] Inpatient medical admissions and inpatient surgical admissions were identified via diagnosis-related groupings (DRG).

**Table S4: Post-matching population sizes by GLP-1RA treatment**

| Members | Full Population | | | | Members with Diabetes | | | | Members without Diabetes | | | |
|---|---|---|---|---|---|---|---|---|---|---|---|---|
| | Treated | | Controls | | Treated | | Controls | | Treated | | Controls | |
| | N | % | N | % | N | % | N | % | N | % | N | % |
| **Overall Population Size** | 742,824 | | 742,824 | | 413,404 | | 413,404 | | 329,420 | | 329,420 | |
| **GLP-1RA Generation (% of Overall Population)** | | | | | | | | | | | | |
| *Semaglutide/Tirzepatide* | 543,097 | 73.1% | | | 278,720 | 67.4% | | | 264,377 | 80.3% | | |
| *Other GLP-1RAs* | 199,727 | 26.9% | | | 134,684 | 32.6% | | | 65,043 | 19.7% | | |

### Table S5: Baseline and follow-up all-cause healthcare costs in matched treated and control cohorts by generation of GLP-1RAs

| Mean (95% confidence interval) of costs ($) PMPM[b] | Full Population | | | Members with Diabetes | | | Members without Diabetes | | |
|---|---|---|---|---|---|---|---|---|---|
| | Baseline | Year 1 | Years 2-6 | Baseline | Year 1 | Years 2-6 | Baseline | Year 1 | Years 2-6 |
| **Total healthcare cost (medical and pharmacy) - including GLP-1RA medications** | | | | | | | | | |
| Treated - Semaglutide or Tirzepatide | $947 ± $5 | $1,819 ± $6 | $1,668 ± $10 | $1,026 ± $7 | $1,911 ± $9 | $1,824 ± $15 | $863 ± $ 7 | $1,721 ± $ 9 | $1,514 ± $ 14 |
| Treated - Other GLP-1RAs | $994 ± $8 | $1,840 ± $11 | $1,816 ± $15 | $1,044 ± $10 | $1,924 ± $14 | $1,950 ± $19 | $890 ± $ 13 | $1,666 ± $ 18 | $1,538 ± $ 23 |
| Controls | $982 ± $4 | $1,082 ± $6 | $1,184 ± $10 | $1,058 ± $6 | $1,188 ± $8 | $1,330 ± $14 | $887 ± $ 6 | $948 ± $ 8 | $1,002 ± $ 12 |
| P-value - Semaglutide or Tirzepatide vs. Controls[c] | <0.0005 | <0.0005 | <0.0005 | <0.0005 | <0.0005 | <0.0005 | <0.0005 | <0.0005 | <0.0005 |
| P-value - Other GLP-1RAs vs. Controls[c] | 0.011 | <0.0005 | <0.0005 | 0.025 | <0.0005 | <0.0005 | 0.695 | <0.0005 | <0.0005 |
| % difference [95% CI] - Semaglutide or Tirzepatide vs. Controls | -3.6% ± 0.7% | 68.1% ± 0.8% | 40.9% ± 1.2% | -3.0% ± 0.9% | 60.8% ± 1.1% | 37.1% ± 1.6% | -2.8% ± 1.0% | 81.7% ± 1.2% | 51.1% ± 1.9% |
| % difference [95% CI] - Other GLP-1RAs vs. Controls | 1.2% ± 0.9% | 70.1% ± 1.2% | 53.4% ± 1.5% | -1.3% ± 1.1% | 61.9% ± 1.4% | 46.6% ± 1.8% | 0.3% ± 1.6% | 75.8% ± 2.0% | 53.4% ± 2.6% |
| **Total healthcare cost (medical and pharmacy) - excluding GLP-1RA medications** | | | | | | | | | |
| Treated - Semaglutide or Tirzepatide | $947 ± $5 | $1,112 ± $6 | $1,168 ± $10 | $1,026 ± $7 | $1,203 ± $9 | $1,267 ± $15 | $863 ± $ 7 | $1,016 ± $ 9 | $1,070 ± $ 14 |
| Treated - Other GLP-1RAs | $994 ± $8 | $1,232 ± $11 | $1,378 ± $15 | $1,044 ± $10 | $1,297 ± $14 | $1,463 ± $19 | $890 ± $ 13 | $1,096 ± $ 17 | $1,202 ± $ 23 |
| Controls | $982 ± $4 | $1,082 ± $6 | $1,184 ± $10 | $1,058 ± $6 | $1,188 ± $8 | $1,330 ± $14 | $887 ± $ 6 | $948 ± $ 8 | $1,002 ± $ 12 |
| P-value - Semaglutide or Tirzepatide vs. Controls[c] | <0.0005 | <0.0005 | 0.029 | <0.0005 | 0.026 | <0.0005 | <0.0005 | <0.0005 | <0.0005 |
| P-value - Other GLP-1RAs vs. Controls[c] | 0.011 | <0.0005 | <0.0005 | 0.025 | <0.0005 | <0.0005 | 0.695 | <0.0005 | <0.0005 |
| % difference [95% CI] - Semaglutide or Tirzepatide vs. Controls | -3.6% ± 0.7% | 2.8% ± 0.8% | -1.3% ± 1.2% | -3.0% ± 0.9% | 1.2% ± 1.1% | -4.8% ± 1.6% | -2.8% ± 1.0% | 7.2% ± 1.2% | 6.8% ± 1.9% |
| % difference [95% CI] - Other GLP-1RAs vs. Controls | 1.2% ± 0.9% | 13.9% ± 1.1% | 16.4% ± 1.5% | -1.3% ± 1.1% | 9.2% ± 1.4% | 10.0% ± 1.8% | 0.3% ± 1.6% | 15.6% ± 2.0% | 20.0% ± 2.6% |
| **Total cost (medical only)** | | | | | | | | | |
| Treated - Semaglutide or Tirzepatide | $635 ± $4 | $764 ± $5 | $801 ± $9 | $652 ± $6 | $795 ± $8 | $838 ± $14 | $618 ± $ 5 | $732 ± $ 7 | $764 ± $ 12 |
| Treated - Other GLP-1RAs | $648 ± $6 | $835 ± $10 | $942 ± $13 | $652 ± $8 | $852 ± $12 | $977 ± $17 | $641 ± $ 10 | $799 ± $ 14 | $871 ± $ 19 |
| Controls | $676 ± $3 | $748 ± $5 | $824 ± $8 | $692 ± $5 | $796 ± $7 | $913 ± $13 | $656 ± $ 5 | $687 ± $ 6 | $713 ± $ 11 |
| P-value - Semaglutide or Tirzepatide vs. Controls[c] | <0.0005 | <0.0005 | <0.0005 | <0.0005 | 0.748 | <0.0005 | <0.0005 | <0.0005 | <0.0005 |
| P-value - Other GLP-1RAs vs. Controls[c] | <0.0005 | <0.0005 | <0.0005 | <0.0005 | <0.0005 | <0.0005 | 0.004 | <0.0005 | <0.0005 |
| % difference [95% CI] - Semaglutide or Tirzepatide vs. Controls | -6.0% ± 0.8% | 2.1% ± 1.0% | -2.8% ± 1.5% | -5.8% ± 1.1% | -0.2% ± 1.4% | -8.3% ± 2.0% | -5.8% ± 1.1% | 6.5% ± 1.4% | 7.2% ± 2.2% |
| % difference [95% CI] - Other GLP-1RAs vs. Controls | -4.1% ± 1.1% | 11.6% ± 1.4% | 14.4% ± 1.9% | -5.7% ± 1.4% | 7.0% ± 1.8% | 7.0% ± 2.3% | -2.4% ± 1.6% | 16.3% ± 2.3% | 22.1% ± 3.1% |

**Table S5: Baseline and follow-up all-cause healthcare costs in matched treated and control cohorts by generation of GLP-1RAs**

| Mean (95% confidence interval) of costs ($) PMPM[b] | Full Population | | | Members with Diabetes | | | Members without Diabetes | | |
|---|---|---|---|---|---|---|---|---|---|
| | Baseline | Year 1 | Years 2-6 | Baseline | Year 1 | Years 2-6 | Baseline | Year 1 | Years 2-6 |
| **Inpatient medical admissions[d]** | | | | | | | | | |
| Treated - Semaglutide or Tirzepatide | $31 ± $1 | $48 ± $2 | $60 ± $3 | $43 ± $1 | $62 ± $3 | $74 ± $5 | $18 ± $1 | $32 ± $2 | $45 ± $4 |
| Treated - Other GLP-1RAs | $42 ± $2 | $68 ± $3 | $106 ± $5 | $52 ± $2 | $82 ± $4 | $128 ± $7 | $20 ± $2 | $41 ± $4 | $60 ± $6 |
| Controls | $33 ± $1 | $66 ± $2 | $91 ± $3 | $46 ± $1 | $86 ± $3 | $124 ± $5 | $17 ± $1 | $41 ± $2 | $49 ± $3 |
| P-value - Semaglutide or Tirzepatide vs. Controls[c] | <0.0005 | <0.0005 | <0.0005 | <0.0005 | <0.0005 | <0.0005 | 0.013 | <0.0005 | 0.092 |
| P-value - Other GLP-1RAs vs. Controls[c] | <0.0005 | 0.189 | <0.0005 | <0.0005 | 0.091 | 0.385 | 0.001 | 0.940 | 0.002 |
| % difference [95% CI] - Semaglutide or Tirzepatide vs. Controls | -7.1% ± 3.6% | -27.9% ± 3.4% | -34.2% ± 4.8% | -7.9% ± 4.1% | -27.5% ± 4.3% | -40.0% ± 5.7% | 9.7% ± 7.6% | -21.9% ± 6.0% | -8.5% ± 9.9% |
| % difference [95% CI] - Other GLP-1RAs vs. Controls | 25.8% ± 5.8% | 3.5% ± 5.3% | 16.7% ± 6.8% | 12.1% ± 5.9% | -4.9% ± 5.7% | 3.2% ± 7.1% | 23.3% ± 13.3% | -0.4% ± 10.3% | 21.3% ± 13.8% |
| **Inpatient surgical admissions[d]** | | | | | | | | | |
| Treated - Semaglutide or Tirzepatide | $58 ± $2 | $89 ± $3 | $98 ± $5 | $75 ± $3 | $105 ± $4 | $116 ± $8 | $40 ± $2 | $73 ± $3 | $80 ± $6 |
| Treated - Other GLP-1RAs | $74 ± $3 | $123 ± $5 | $142 ± $7 | $90 ± $4 | $137 ± $7 | $161 ± $9 | $40 ± $4 | $96 ± $7 | $101 ± $9 |
| Controls | $60 ± $2 | $103 ± $3 | $120 ± $5 | $80 ± $2 | $127 ± $4 | $155 ± $8 | $36 ± $2 | $73 ± $3 | $77 ± $6 |
| P-value - Semaglutide or Tirzepatide vs. Controls[c] | 0.021 | <0.0005 | <0.0005 | 0.005 | <0.0005 | <0.0005 | 0.004 | 0.967 | 0.514 |
| P-value - Other GLP-1RAs vs. Controls[c] | <0.0005 | <0.0005 | <0.0005 | <0.0005 | 0.010 | 0.327 | 0.034 | <0.0005 | <0.0005 |
| % difference [95% CI] - Semaglutide or Tirzepatide vs. Controls | -4.5% ± 3.8% | -12.9% ± 3.5% | -18.9% ± 5.9% | -6.5% ± 4.6% | -16.8% ± 4.4% | -25.5% ± 7.0% | 10.8% ± 7.3% | 0.1% ± 6.0% | 3.7% ± 11.1% |
| % difference [95% CI] - Other GLP-1RAs vs. Controls | 23.1% ± 6.0% | 20.1% ± 5.5% | 17.8% ± 7.0% | 13.5% ± 6.3% | 8.0% ± 6.1% | 3.8% ± 7.6% | 12.8% ± 11.9% | 31.8% ± 10.4% | 31.5% ± 14.4% |
| **Observation stays** | | | | | | | | | |
| Treated - Semaglutide or Tirzepatide | $26 ± $1 | $31 ± $1 | $29 ± $1 | $29 ± $1 | $34 ± $1 | $30 ± $2 | $24 ± $1 | $28 ± $1 | $27 ± $2 |
| Treated - Other GLP-1RAs | $30 ± $1 | $35 ± $1 | $36 ± $2 | $32 ± $1 | $37 ± $2 | $37 ± $2 | $27 ± $2 | $31 ± $2 | $33 ± $3 |
| Controls | $36 ± $1 | $30 ± $1 | $29 ± $1 | $37 ± $1 | $34 ± $1 | $31 ± $1 | $35 ± $1 | $25 ± $1 | $26 ± $2 |
| P-value - Semaglutide or Tirzepatide vs. Controls[c] | <0.0005 | 0.123 | 0.674 | <0.0005 | 0.988 | 0.168 | <0.0005 | 0.001 | 0.252 |
| P-value - Other GLP-1RAs vs. Controls[c] | <0.0005 | <0.0005 | <0.0005 | <0.0005 | 0.001 | <0.0005 | <0.0005 | <0.0005 | <0.0005 |
| % difference [95% CI] - Semaglutide or Tirzepatide vs. Controls | -26.8% ± 2.6% | 2.7% ± 3.4% | -1.2% ± 5.7% | -22.5% ± 3.6% | -0.0% ± 4.4% | -4.9% ± 6.9% | -31.1% ± 3.9% | 9.3% ± 5.4% | 5.7% ± 9.7% |
| % difference [95% CI] - Other GLP-1RAs vs. Controls | -16.6% ± 3.8% | 16.8% ± 4.8% | 24.4% ± 6.7% | -15.2% ± 4.7% | 9.7% ± 5.6% | 18.5% ± 7.6% | -22.1% ± 6.2% | 22.5% ± 8.9% | 29.0% ± 13.1% |

**Table S5: Baseline and follow-up all-cause healthcare costs in matched treated and control cohorts by generation of GLP-1RAs**

| Mean (95% confidence interval) of costs ($) PMPM[b] | Full Population | | | Members with Diabetes | | | Members without Diabetes | | |
|---|---|---|---|---|---|---|---|---|---|
| | Baseline | Year 1 | Years 2-6 | Baseline | Year 1 | Years 2-6 | Baseline | Year 1 | Years 2-6 |
| **Emergency department visits** | | | | | | | | | |
| Treated - Semaglutide or Tirzepatide | $53 ± $1 | $59 ± $1 | $59 ± $1 | $53 ± $1 | $58 ± $1 | $57 ± $1 | $52 ± $1 | $61 ± $1 | $60 ± $2 |
| Treated - Other GLP-1RAs | $55 ± $1 | $61 ± $1 | $61 ± $1 | $55 ± $1 | $60 ± $2 | $60 ± $2 | $55 ± $2 | $63 ± $2 | $63 ± $2 |
| Controls | $62 ± $1 | $60 ± $1 | $58 ± $1 | $62 ± $1 | $60 ± $1 | $59 ± $1 | $64 ± $1 | $59 ± $1 | $57 ± $1 |
| P-value - Semaglutide or Tirzepatide vs. Controls[c] | <0.0005 | 0.392 | 0.454 | <0.0005 | <0.0005 | 0.013 | <0.0005 | 0.002 | 0.001 |
| P-value - Other GLP-1RAs vs. Controls[c] | <0.0005 | 0.034 | 0.001 | <0.0005 | 0.966 | 0.499 | <0.0005 | <0.0005 | <0.0005 |
| % difference [95% CI] - Semaglutide or Tirzepatide vs. Controls | -15.7% ± 1.2% | -0.6% ± 1.4% | 0.9% ± 2.4% | -13.2% ± 1.7% | -3.9% ± 2.0% | -3.8% ± 3.0% | -18.4% ± 1.7% | 3.2% ± 2.0% | 6.3% ± 3.8% |
| % difference [95% CI] - Other GLP-1RAs vs. Controls | -11.9% ± 1.9% | 2.5% ± 2.3% | 4.6% ± 2.7% | -10.8% ± 2.5% | -0.1% ± 3.0% | 1.2% ± 3.4% | -13.1% ± 3.0% | 6.8% ± 3.4% | 10.3% ± 4.7% |
| **Ambulatory services - provider visits** | | | | | | | | | |
| Treated - Semaglutide or Tirzepatide | $105 ± $0 | $115 ± $0 | $109 ± $1 | $100 ± $1 | $111 ± $1 | $105 ± $1 | $111 ± $1 | $120 ± $1 | $114 ± $1 |
| Treated - Other GLP-1RAs | $103 ± $1 | $116 ± $1 | $113 ± $1 | $95 ± $1 | $110 ± $1 | $107 ± $1 | $119 ± $1 | $128 ± $1 | $126 ± $2 |
| Controls | $100 ± $0 | $98 ± $0 | $99 ± $0 | $96 ± $0 | $95 ± $1 | $95 ± $1 | $106 ± $1 | $103 ± $1 | $102 ± $1 |
| P-value - Semaglutide or Tirzepatide vs. Controls[c] | <0.0005 | <0.0005 | <0.0005 | <0.0005 | <0.0005 | <0.0005 | <0.0005 | <0.0005 | <0.0005 |
| P-value - Other GLP-1RAs vs. Controls[c] | <0.0005 | <0.0005 | <0.0005 | 0.042 | <0.0005 | <0.0005 | <0.0005 | <0.0005 | <0.0005 |
| % difference [95% CI] - Semaglutide or Tirzepatide vs. Controls | 4.6% ± 0.5% | 17.1% ± 0.6% | 11.0% ± 0.8% | 3.6% ± 0.7% | 16.6% ± 0.8% | 9.8% ± 1.0% | 4.7% ± 0.8% | 16.8% ± 0.9% | 11.3% ± 1.2% |
| % difference [95% CI] - Other GLP-1RAs vs. Controls | 2.5% ± 0.8% | 17.6% ± 0.8% | 14.7% ± 1.1% | -1.0% ± 1.0% | 15.5% ± 1.0% | 11.8% ± 1.5% | 12.4% ± 1.3% | 25.0% ± 1.4% | 23.3% ± 1.7% |

**Table S5: Baseline and follow-up all-cause healthcare costs in matched treated and control cohorts by generation of GLP-1RAs**

| Mean (95% confidence interval) of costs ($) PMPM[b] | Full Population | | | Members with Diabetes | | | Members without Diabetes | | |
|---|---|---|---|---|---|---|---|---|---|
| | Baseline | Year 1 | Years 2-6 | Baseline | Year 1 | Years 2-6 | Baseline | Year 1 | Years 2-6 |
| **Ambulatory services - general laboratory testing** | | | | | | | | | |
| Treated - Semaglutide or Tirzepatide | $27 ± $0 | $25 ± $0 | $26 ± $0 | $28 ± $0 | $27 ± $0 | $28 ± $0 | $26 ± $0 | $24 ± $0 | $25 ± $0 |
| Treated - Other GLP-1RAs | $27 ± $0 | $27 ± $0 | $28 ± $0 | $27 ± $0 | $27 ± $0 | $28 ± $0 | $27 ± $0 | $26 ± $1 | $27 ± $0 |
| Controls | $23 ± $0 | $22 ± $0 | $23 ± $0 | $25 ± $0 | $24 ± $0 | $25 ± $0 | $21 ± $0 | $20 ± $0 | $20 ± $0 |
| P-value - Semaglutide or Tirzepatide vs. Controls[c] | <0.0005 | <0.0005 | <0.0005 | <0.0005 | <0.0005 | <0.0005 | <0.0005 | <0.0005 | <0.0005 |
| P-value - Other GLP-1RAs vs. Controls[c] | <0.0005 | <0.0005 | <0.0005 | <0.0005 | <0.0005 | <0.0005 | <0.0005 | <0.0005 | <0.0005 |
| % difference [95% CI] - Semaglutide or Tirzepatide vs. Controls | 14.4% ± 0.7% | 13.3% ± 0.8% | 14.7% ± 1.3% | 8.9% ± 0.9% | 10.4% ± 1.0% | 9.7% ± 1.7% | 23.9% ± 1.1% | 19.3% ± 1.2% | 24.3% ± 1.9% |
| % difference [95% CI] - Other GLP-1RAs vs. Controls | 17.3% ± 1.1% | 19.4% ± 1.2% | 20.8% ± 1.5% | 7.6% ± 1.2% | 11.4% ± 1.3% | 11.8% ± 1.7% | 32.5% ± 2.1% | 30.1% ± 2.8% | 32.8% ± 2.7% |
| **Ambulatory services - imaging** | | | | | | | | | |
| Treated - Semaglutide or Tirzepatide | $57 ± $0 | $63 ± $0 | $67 ± $1 | $54 ± $0 | $62 ± $1 | $65 ± $1 | $60 ± $0 | $65 ± $1 | $69 ± $1 |
| Treated - Other GLP-1RAs | $54 ± $1 | $63 ± $1 | $67 ± $1 | $50 ± $1 | $61 ± $1 | $65 ± $1 | $62 ± $1 | $67 ± $1 | $72 ± $1 |
| Controls | $56 ± $0 | $58 ± $0 | $62 ± $1 | $54 ± $0 | $56 ± $0 | $61 ± $1 | $59 ± $0 | $60 ± $0 | $63 ± $1 |
| P-value - Semaglutide or Tirzepatide vs. Controls[c] | <0.0005 | <0.0005 | <0.0005 | 0.019 | <0.0005 | <0.0005 | <0.0005 | <0.0005 | <0.0005 |
| P-value - Other GLP-1RAs vs. Controls[c] | <0.0005 | <0.0005 | <0.0005 | <0.0005 | <0.0005 | <0.0005 | <0.0005 | <0.0005 | <0.0005 |
| % difference [95% CI] - Semaglutide or Tirzepatide vs. Controls | 2.2% ± 0.8% | 9.2% ± 0.9% | 8.9% ± 1.4% | 1.3% ± 1.1% | 10.3% ± 1.3% | 6.5% ± 1.9% | 2.2% ± 1.1% | 7.6% ± 1.2% | 10.9% ± 2.0% |
| % difference [95% CI] - Other GLP-1RAs vs. Controls | -3.9% ± 1.1% | 8.3% ± 1.3% | 9.4% ± 1.5% | -7.0% ± 1.3% | 7.9% ± 1.8% | 7.1% ± 2.0% | 5.3% ± 1.8% | 11.6% ± 1.9% | 15.1% ± 2.5% |
| **Ambulatory services - dialysis** | | | | | | | | | |
| Treated - Semaglutide or Tirzepatide | $0 ± $0 | $1 ± $0 | $3 ± $1 | $0 ± $0 | $1 ± $0 | $6 ± $1 | $0 ± $0 | $0 ± $0 | $0 ± $0 |
| Treated - Other GLP-1RAs | $0 ± $0 | $1 ± $0 | $8 ± $1 | $0 ± $0 | $1 ± $0 | $12 ± $2 | $0 ± $0 | $0 ± $0 | $1 ± $1 |
| Controls | $0 ± $0 | $2 ± $0 | $9 ± $1 | $0 ± $0 | $4 ± $1 | $15 ± $2 | $0 ± $0 | $0 ± $0 | $1 ± $1 |
| P-value - Semaglutide or Tirzepatide vs. Controls[c] | - | <0.0005 | <0.0005 | - | <0.0005 | <0.0005 | - | 0.006 | 0.008 |
| P-value - Other GLP-1RAs vs. Controls[c] | - | <0.0005 | 0.351 | - | <0.0005 | 0.008 | - | 0.308 | 0.304 |
| % difference [95% CI] - Semaglutide or Tirzepatide vs. Controls | - | -66.0% ± 18.9% | -64.1% ± 12.4% | - | -61.7% ± 20.3% | -59.8% ± 13.3% | - | -85.2% ± 60.9% | -72.8% ± 53.6% |
| % difference [95% CI] - Other GLP-1RAs vs. Controls | - | -54.0% ± 21.8% | -8.8% ± 18.4% | - | -61.5% ± 20.9% | -22.7% ± 16.8% | - | -47.5% ± 91.4% | -36.9% ± 70.4% |

**Table S5: Baseline and follow-up all-cause healthcare costs in matched treated and control cohorts by generation of GLP-1RAs**

| Mean (95% confidence interval) of costs ($) PMPM[b] | Full Population | | | Members with Diabetes | | | Members without Diabetes | | |
|---|---|---|---|---|---|---|---|---|---|
| | Baseline | Year 1 | Years 2-6 | Baseline | Year 1 | Years 2-6 | Baseline | Year 1 | Years 2-6 |
| **Ambulatory services - other services** | | | | | | | | | |
| Treated - Semaglutide or Tirzepatide | $279 ± $2 | $332 ± $3 | $350 ± $5 | $271 ± $3 | $335 ± $4 | $357 ± $7 | $288 ± $ 4 | $330 ± $ 4 | $343 ± $ 7 |
| Treated - Other GLP-1RAs | $264 ± $4 | $341 ± $5 | $382 ± $6 | $251 ± $4 | $338 ± $6 | $379 ± $8 | $289 ± $ 6 | $347 ± $ 8 | $389 ± $ 12 |
| Controls | $305 ± $2 | $309 ± $3 | $334 ± $4 | $292 ± $3 | $311 ± $4 | $346 ± $6 | $321 ± $ 3 | $306 ± $ 4 | $318 ± $ 6 |
| P-value - Semaglutide or Tirzepatide vs. Controls[c] | <0.0005 | <0.0005 | <0.0005 | <0.0005 | <0.0005 | 0.016 | <0.0005 | <0.0005 | <0.0005 |
| P-value - Other GLP-1RAs vs. Controls[c] | <0.0005 | <0.0005 | <0.0005 | <0.0005 | <0.0005 | <0.0005 | <0.0005 | <0.0005 | <0.0005 |
| % difference [95% CI] - Semaglutide or Tirzepatide vs. Controls | -8.3% ± 1.1% | 7.6% ± 1.3% | 5.0% ± 2.0% | -7.2% ± 1.6% | 7.6% ± 1.8% | 3.2% ± 2.6% | -10.2% ± 1.5% | 7.8% ± 1.9% | 7.9% ± 3.0% |
| % difference [95% CI] - Other GLP-1RAs vs. Controls | -13.4% ± 1.4% | 10.3% ± 1.8% | 14.4% ± 2.3% | -13.9% ± 1.8% | 8.6% ± 2.3% | 9.4% ± 2.8% | -9.7% ± 2.3% | 13.4% ± 3.0% | 22.1% ± 4.2% |
| **GLP-1RAs** | | | | | | | | | |
| Treated - Semaglutide or Tirzepatide | $0 ± $0 | $707 ± $1 | $500 ± $2 | $0 ± $0 | $708 ± $1 | $557 ± $2 | $0 ± $0 | $706 ± $2 | $444 ± $2 |
| Treated - Other GLP-1RAs | $0 ± $0 | $608 ± $2 | $438 ± $2 | $0 ± $0 | $627 ± $2 | $487 ± $2 | $0 ± $0 | $570 ± $3 | $336 ± $4 |
| Controls | $0 ± $0 | - | - | $0 ± $0 | - | - | $0 ± $0 | - | - |
| P-value - Semaglutide or Tirzepatide vs. Controls[c] | - | - | - | - | - | - | - | - | - |
| P-value - Other GLP-1RAs vs. Controls[c] | - | - | - | - | - | - | - | - | - |
| % difference [95% CI] - Semaglutide or Tirzepatide vs. Controls | - | - | - | - | - | - | - | - | - |
| % difference [95% CI] - Other GLP-1RAs vs. Controls | - | - | - | - | - | - | - | - | - |
| **Pharmaceuticals excluding GLP-1RAs** | | | | | | | | | |
| Treated - Semaglutide or Tirzepatide | $311 ± $3 | $348 ± $3 | $368 ± $5 | $375 ± $4 | $408 ± $4 | $429 ± $6 | $245 ± $4 | $284 ± $5 | $306 ± $7 |
| Treated - Other GLP-1RAs | $346 ± $4 | $397 ± $5 | $436 ± $6 | $392 ± $5 | $446 ± $5 | $486 ± $7 | $250 ± $8 | $297 ± $9 | $332 ± $11 |
| Controls | $306 ± $3 | $334 ± $3 | $360 ± $4 | $366 ± $3 | $392 ± $4 | $417 ± $5 | $231 ± $4 | $260 ± $4 | $289 ± $6 |
| P-value - Semaglutide or Tirzepatide vs. Controls[c] | 0.009 | <0.0005 | 0.017 | 0.002 | <0.0005 | 0.003 | <0.0005 | <0.0005 | <0.0005 |
| P-value - Other GLP-1RAs vs. Controls[c] | <0.0005 | <0.0005 | <0.0005 | <0.0005 | <0.0005 | <0.0005 | <0.0005 | <0.0005 | <0.0005 |
| % difference [95% CI] - Semaglutide or Tirzepatide vs. Controls | 1.7% ± 1.2% | 4.2% ± 1.3% | 2.1% ± 1.7% | 2.3% ± 1.4% | 4.1% ± 1.5% | 2.9% ± 1.9% | 5.9% ± 2.4% | 9.0% ± 2.4% | 5.8% ± 3.2% |
| % difference [95% CI] - Other GLP-1RAs vs. Controls | 12.9% ± 1.6% | 19.0% ± 1.6% | 20.9% ± 1.9% | 7.1% ± 1.6% | 13.7% ± 1.7% | 16.4% ± 2.0% | 8.0% ± 3.7% | 13.9% ± 3.7% | 14.6% ± 4.2% |

[a] Members included in this table were required to have at least 12 months of baseline and at least 6 months of follow-up continuous enrollment. Utilization is assigned to the time period based on the start date of the stay, procedure, or prescription fill.

[b] All costs were adjusted using the medical component of the Consumer Price Index for Urban Consumers to reflect inflation of costs between the time when medical care was provided and the end date of the current analysis (August 31, 2024). Costs were capped at $500,000 per year and scaled downward if the total medical and pharmacy costs exceeded that amount.

[c] Mean costs were compared with the Student's T Test. Without a Bonferroni correction for the 2 statistical tests done within each utilization category, population subgroup, and time period, a p-value less than 0.05 denotes a statistically significant difference, and a p-value less than 0.001 is a highly statistically significant difference. With a Bonferroni correction, only p-values less than 0.025 denote statistically significant differences, and p-values less than 0.0005 denote highly statistically significant differences.

[d] Inpatient medical admissions and inpatient surgical admissions were identified via diagnosis-related groupings (DRG).

**Table S6: Follow-up gastrointestinal side effect-related medical and pharmacy costs in matched treated and control cohorts,[a] by index GLP-1RA type**

| Mean (95% confidence interval) of costs ($) PMPM[b] | Full Population | | Members with Diabetes | | Members without Diabetes | |
|---|---|---|---|---|---|---|
| | Year 1 | Years 2-6 | Year 1 | Years 2-6 | Year 1 | Years 2-6 |
| **Total cost (medical only)** | | | | | | |
| Treated - Semaglutide or Tirzepatide | $126 ± $ 2 | $127 ± $ 3 | $120 ± $ 3 | $118 ± $ 4 | $133 ± $ 3 | $136 ± $ 5 |
| Treated - Other GLP-1RAs | $136 ± $ 3 | $140 ± $ 4 | $127 ± $ 4 | $135 ± $ 5 | $153 ± $ 5 | $153 ± $ 6 |
| Controls | $116 ± $ 2 | $121 ± $ 3 | $117 ± $ 2 | $128 ± $ 5 | $114 ± $ 2 | $113 ± $ 3 |
| P-value - Semaglutide or Tirzepatide vs. Controls[c] | <0.0005 | 0.0152 | 0.1046 | 0.0048 | <0.0005 | <0.0005 |
| P-value - Other GLP-1RAs vs. Controls[c] | <0.0005 | <0.0005 | <0.0005 | 0.0708 | <0.0005 | <0.0005 |
| % difference [95% CI] - Semaglutide or Tirzepatide vs. Controls | 9.0% ± 2.2% | 4.7% ± 3.8% | 2.5% ± 3.0% | -7.5% ± 5.2% | 16.4% ± 3.2% | 19.9% ± 5.3% |
| % difference [95% CI] - Other GLP-1RAs vs. Controls | 17.3% ± 3.2% | 15.6% ± 4.1% | 8.7% ± 4.2% | 5.0% ± 5.5% | 34.3% ± 5.2% | 34.8% ± 6.3% |
| **Inpatient medical admissions[d]** | | | | | | |
| Treated - Semaglutide or Tirzepatide | $12 ± $ 1 | $12 ± $ 1 | $14 ± $ 1 | $14 ± $ 1 | $10 ± $ 1 | $11 ± $ 1 |
| Treated - Other GLP-1RAs | $16 ± $ 2 | $21 ± $ 2 | $19 ± $ 2 | $24 ± $ 2 | $12 ± $ 2 | $15 ± $ 3 |
| Controls | $14 ± $ 1 | $17 ± $ 1 | $17 ± $ 1 | $22 ± $ 2 | $10 ± $ 1 | $11 ± $ 1 |
| P-value - Semaglutide or Tirzepatide vs. Controls[c] | <0.0005 | <0.0005 | <0.0005 | <0.0005 | 0.8809 | 0.7844 |
| P-value - Other GLP-1RAs vs. Controls[c] | 0.0065 | 0.0026 | 0.1692 | 0.3888 | 0.1538 | 0.0271 |
| % difference [95% CI] - Semaglutide or Tirzepatide vs. Controls | -13.4% ± 6.4% | -28.5% ± 8.8% | -18.7% ± 7.3% | -38.2% ± 10.2% | 1.0% ± 12.7% | -2.4% ± 17.4% |
| % difference [95% CI] - Other GLP-1RAs vs. Controls | 16.9% ± 12.2% | 18.9% ± 12.3% | 9.8% ± 13.9% | 5.8% ± 13.1% | 13.7% ± 18.8% | 30.2% ± 26.7% |

**Table S6: Follow-up gastrointestinal side effect-related medical and pharmacy costs in matched treated and control cohorts,[a] by index GLP-1RA type**

| Mean (95% confidence interval) of costs ($) PMPM[b] | Full Population | | Members with Diabetes | | Members without Diabetes | |
|---|---|---|---|---|---|---|
| | Year 1 | Years 2-6 | Year 1 | Years 2-6 | Year 1 | Years 2-6 |
| **Inpatient surgical admissions[d]** | | | | | | |
| Treated - Semaglutide or Tirzepatide | $23 ± $ 1 | $25 ± $ 2 | $22 ± $ 2 | $23 ± $ 2 | $24 ± $ 2 | $28 ± $ 3 |
| Treated - Other GLP-1RAs | $28 ± $ 2 | $28 ± $ 2 | $26 ± $ 2 | $28 ± $ 2 | $32 ± $ 3 | $28 ± $ 3 |
| Controls | $23 ± $ 1 | $25 ± $ 3 | $26 ± $ 2 | $30 ± $ 5 | $19 ± $ 1 | $18 ± $ 2 |
| P-value - Semaglutide or Tirzepatide vs. Controls[c] | 0.5963 | 0.5935 | 0.0024 | 0.0088 | <0.0005 | <0.0005 |
| P-value - Other GLP-1RAs vs. Controls[c] | <0.0005 | 0.0533 | 0.8932 | 0.4470 | <0.0005 | <0.0005 |
| % difference [95% CI] - Semaglutide or Tirzepatide vs. Controls | 1.9% ± 7.0% | 3.7% ± 13.4% | -13.5% ± 8.7% | -22.8% ± 17.0% | 26.7% ± 11.5% | 53.5% ± 20.5% |
| % difference [95% CI] - Other GLP-1RAs vs. Controls | 23.1% ± 9.7% | 12.9% ± 13.1% | 0.8% ± 11.4% | -6.7% ± 17.2% | 69.9% ± 18.1% | 52.5% ± 18.7% |
| **Observation stays** | | | | | | |
| Treated - Semaglutide or Tirzepatide | $12 ± $0 | $11 ± $1 | $11 ± $ 1 | $10 ± $ 1 | $12 ± $ 1 | $11 ± $ 1 |
| Treated - Other GLP-1RAs | $12 ± $1 | $13 ± $1 | $11 ± $ 1 | $12 ± $ 1 | $14 ± $ 1 | $15 ± $ 2 |
| Controls | $11 ± $0 | $10 ± $1 | $11 ± $ 1 | $10 ± $ 1 | $10 ± $ 1 | $10 ± $ 1 |
| P-value - Semaglutide or Tirzepatide vs. Controls[c] | <0.0005 | 0.2430 | 0.6468 | 0.6548 | <0.0005 | 0.0441 |
| P-value - Other GLP-1RAs vs. Controls[c] | <0.0005 | <0.0005 | 0.5583 | 0.0158 | <0.0005 | <0.0005 |
| % difference [95% CI] - Semaglutide or Tirzepatide vs. Controls | 11.4% ± 5.8% | 5.7% ± 9.6% | 1.7% ± 7.5% | -2.8% ± 12.3% | 23.4% ± 9.2% | 15.4% ± 15.0% |
| % difference [95% CI] - Other GLP-1RAs vs. Controls | 16.1% ± 8.1% | 27.4% ± 10.3% | 2.9% ± 9.6% | 14.4% ± 11.7% | 42.3% ± 15.2% | 53.0% ± 20.4% |
| **Emergency department visits** | | | | | | |
| Treated - Semaglutide or Tirzepatide | $21 ± $0 | $19 ± $1 | $19 ± $0 | $17 ± $1 | $24 ± $1 | $21 ± $1 |
| Treated - Other GLP-1RAs | $20 ± $1 | $19 ± $1 | $18 ± $1 | $17 ± $1 | $25 ± $1 | $23 ± $1 |
| Controls | $18 ± $0 | $17 ± $0 | $17 ± $1 | $16 ± $1 | $19 ± $0 | $18 ± $1 |
| P-value - Semaglutide or Tirzepatide vs. Controls[c] | <0.0005 | <0.0005 | <0.0005 | 0.0224 | <0.0005 | <0.0005 |
| P-value - Other GLP-1RAs vs. Controls[c] | <0.0005 | 0.0007 | 0.0180 | 0.3911 | <0.0005 | <0.0005 |
| % difference [95% CI] - Semaglutide or Tirzepatide vs. Controls | 19.5% ± 2.8% | 10.8% ± 4.2% | 12.6% ± 4.1% | 6.6% ± 5.7% | 25.2% ± 3.6% | 13.0% ± 6.1% |
| % difference [95% CI] - Other GLP-1RAs vs. Controls | 13.7% ± 3.9% | 7.8% ± 4.5% | 6.2% ± 5.1% | 2.3% ± 5.3% | 31.7% ± 6.5% | 22.6% ± 8.6% |

**Table S6: Follow-up gastrointestinal side effect-related medical and pharmacy costs in matched treated and control cohorts,[a] by index GLP-1RA type**

| Mean (95% confidence interval) of costs ($) PMPM[b] | Full Population | | Members with Diabetes | | Members without Diabetes | |
|---|---|---|---|---|---|---|
| | Year 1 | Years 2-6 | Year 1 | Years 2-6 | Year 1 | Years 2-6 |
| **Ambulatory services - any** | | | | | | |
| Treated - Semaglutide or Tirzepatide | $49 ± $ 1 | $52 ± $ 2 | $46 ± $ 1 | $47 ± $ 2 | $53 ± $ 1 | $56 ± $ 2 |
| Treated - Other GLP-1RAs | $48 ± $ 1 | $52 ± $ 2 | $44 ± $ 2 | $47 ± $ 2 | $58 ± $ 2 | $62 ± $ 3 |
| Controls | $43 ± $ 1 | $45 ± $ 1 | $39 ± $ 1 | $43 ± $ 1 | $47 ± $ 1 | $48 ± $ 2 |
| P-value - Semaglutide or Tirzepatide vs. Controls[c] | <0.0005 | <0.0005 | <0.0005 | <0.0005 | <0.0005 | <0.0005 |
| P-value - Other GLP-1RAs vs. Controls[c] | <0.0005 | <0.0005 | <0.0005 | <0.0005 | <0.0005 | <0.0005 |
| % difference [95% CI] - Semaglutide or Tirzepatide vs. Controls | 16.0% ± 2.5% | 14.4% ± 4.2% | 17.8% ± 3.8% | 11.1% ± 6.0% | 12.6% ± 3.4% | 15.8% ± 5.8% |
| % difference [95% CI] - Other GLP-1RAs vs. Controls | 13.0% ± 3.5% | 15.0% ± 4.5% | 11.8% ± 4.6% | 10.5% ± 5.7% | 22.2% ± 5.6% | 28.4% ± 7.6% |
| **Ambulatory services - evaluation and management** | | | | | | |
| Treated - Semaglutide or Tirzepatide | $7 ± $0 | $6 ± $0 | $6 ± $0 | $5 ± $0 | $7 ± $0 | $7 ± $0 |
| Treated - Other GLP-1RAs | $6 ± $0 | $6 ± $0 | $6 ± $0 | $5 ± $0 | $8 ± $0 | $7 ± $0 |
| Controls | $5 ± $0 | $5 ± $0 | $5 ± $0 | $5 ± $0 | $6 ± $0 | $6 ± $0 |
| P-value - Semaglutide or Tirzepatide vs. Controls[c] | <0.0005 | <0.0005 | <0.0005 | <0.0005 | <0.0005 | <0.0005 |
| P-value - Other GLP-1RAs vs. Controls[c] | <0.0005 | <0.0005 | <0.0005 | <0.0005 | <0.0005 | <0.0005 |
| % difference [95% CI] - Semaglutide or Tirzepatide vs. Controls | 21.6% ± 1.1% | 10.7% ± 1.6% | 20.4% ± 1.6% | 8.6% ± 2.4% | 20.2% ± 1.5% | 10.0% ± 2.2% |
| % difference [95% CI] - Other GLP-1RAs vs. Controls | 16.5% ± 1.5% | 7.5% ± 1.8% | 14.7% ± 2.0% | 5.6% ± 2.3% | 28.5% ± 2.6% | 18.6% ± 3.1% |
| **Ambulatory services - general laboratory testing** | | | | | | |
| Treated - Semaglutide or Tirzepatide | $2 ± $0 | $2 ± $0 | $1 ± $0 | $2 ± $0 | $2 ± $0 | $2 ± $0 |
| Treated - Other GLP-1RAs | $2 ± $0 | $2 ± $0 | $1 ± $0 | $1 ± $0 | $2 ± $0 | $2 ± $0 |
| Controls | $1 ± $0 | $1 ± $0 | $1 ± $0 | $1 ± $0 | $1 ± $0 | $1 ± $0 |
| P-value - Semaglutide or Tirzepatide vs. Controls[c] | <0.0005 | <0.0005 | <0.0005 | <0.0005 | <0.0005 | <0.0005 |
| P-value - Other GLP-1RAs vs. Controls[c] | <0.0005 | <0.0005 | <0.0005 | <0.0005 | <0.0005 | <0.0005 |
| % difference [95% CI] - Semaglutide or Tirzepatide vs. Controls | 15.4% ± 2.8% | 16.7% ± 4.7% | 15.3% ± 4.0% | 13.2% ± 6.3% | 14.6% ± 3.8% | 19.2% ± 7.0% |
| % difference [95% CI] - Other GLP-1RAs vs. Controls | 17.2% ± 3.5% | 16.0% ± 4.8% | 13.8% ± 4.4% | 11.3% ± 6.1% | 27.2% ± 5.9% | 27.8% ± 8.3% |
| **Ambulatory services - imaging** | | | | | | |
| Treated - Semaglutide or Tirzepatide | $5 ± $0 | $5 ± $0 | $4 ± $0 | $4 ± $0 | $5 ± $0 | $5 ± $0 |
| Treated - Other GLP-1RAs | $5 ± $0 | $5 ± $0 | $4 ± $0 | $5 ± $0 | $6 ± $0 | $6 ± $0 |
| Controls | $4 ± $0 | $4 ± $0 | $4 ± $0 | $4 ± $0 | $5 ± $0 | $5 ± $0 |
| P-value - Semaglutide or Tirzepatide vs. Controls[c] | <0.0005 | 0.0028 | <0.0005 | 0.0331 | 0.0055 | 0.1023 |
| P-value - Other GLP-1RAs vs. Controls[c] | <0.0005 | <0.0005 | <0.0005 | <0.0005 | <0.0005 | <0.0005 |
| % difference [95% CI] - Semaglutide or Tirzepatide vs. Controls | 10.7% ± 3.1% | 7.1% ± 4.6% | 14.7% ± 4.8% | 7.0% ± 6.4% | 5.6% ± 4.0% | 5.4% ± 6.5% |
| % difference [95% CI] - Other GLP-1RAs vs. Controls | 15.9% ± 4.0% | 15.5% ± 4.9% | 13.9% ± 5.3% | 15.1% ± 6.6% | 26.1% ± 6.4% | 22.4% ± 7.8% |

**Table S6: Follow-up gastrointestinal side effect-related medical and pharmacy costs in matched treated and control cohorts,[a] by index GLP-1RA type**

| Mean (95% confidence interval) of costs ($) PMPM[b] | Full Population | | Members with Diabetes | | Members without Diabetes | |
|---|---|---|---|---|---|---|
| | Year 1 | Years 2-6 | Year 1 | Years 2-6 | Year 1 | Years 2-6 |
| **Ambulatory services - other services** | | | | | | |
| Treated - Semaglutide or Tirzepatide | $37 ± $ 1 | $39 ± $ 1 | $34 ± $ 1 | $36 ± $ 2 | $39 ± $ 1 | $43 ± $ 2 |
| Treated - Other GLP-1RAs | $36 ± $ 1 | $39 ± $ 2 | $32 ± $ 2 | $36 ± $ 2 | $42 ± $ 2 | $47 ± $ 3 |
| Controls | $32 ± $ 1 | $34 ± $ 1 | $29 ± $ 1 | $32 ± $ 1 | $35 ± $ 1 | $36 ± $ 2 |
| P-value - Semaglutide or Tirzepatide vs. Controls[c] | <0.0005 | <0.0005 | <0.0005 | 0.0015 | <0.0005 | <0.0005 |
| P-value - Other GLP-1RAs vs. Controls[c] | <0.0005 | <0.0005 | <0.0005 | 0.0040 | <0.0005 | <0.0005 |
| % difference [95% CI] - Semaglutide or Tirzepatide vs. Controls | 15.8% ± 3.3% | 16.1% ± 5.3% | 18.1% ± 4.9% | 12.4% ± 7.6% | 12.1% ± 4.3% | 18.1% ± 7.4% |
| % difference [95% CI] - Other GLP-1RAs vs. Controls | 11.9% ± 4.5% | 16.1% ± 5.7% | 11.1% ± 5.9% | 10.6% ± 7.2% | 20.5% ± 7.2% | 30.8% ± 9.7% |
| **Pharmaceuticals for the management of gastrointestinal side-effects** | | | | | | |
| Treated - Semaglutide or Tirzepatide | $3 ± $0 | $4 ± $0 | $2 ± $0 | $3 ± $1 | $3 ± $0 | $4 ± $0 |
| Treated - Other GLP-1RAs | $3 ± $0 | $4 ± $0 | $2 ± $0 | $3 ± $0 | $6 ± $1 | $6 ± $1 |
| Controls | $3 ± $1 | $4 ± $0 | $2 ± $0 | $3 ± $0 | $4 ± $1 | $4 ± $1 |
| P-value - Semaglutide or Tirzepatide vs. Controls[c] | 0.3520 | 0.7296 | <0.0005 | 0.6017 | 0.7709 | 0.2122 |
| P-value - Other GLP-1RAs vs. Controls[c] | 0.0106 | 0.0322 | <0.0005 | 0.3957 | 0.0126 | 0.0029 |
| % difference [95% CI] - Semaglutide or Tirzepatide vs. Controls | 9.9% ± 20.9% | -3.0% ± 17.1% | 28.5% ± 13.6% | 7.6% ± 28.6% | -4.9% ± 33.2% | -13.6% ± 21.4% |
| % difference [95% CI] - Other GLP-1RAs vs. Controls | 31.3% ± 24.0% | 18.0% ± 16.4% | 31.3% ± 14.1% | 8.1% ± 18.8% | 54.2% ± 42.6% | 45.1% ± 29.6% |

a Members included in this table were required to have at least 12 months of baseline and at least 6 months of follow-up continuous enrollment. Gastrointestinal side-effect related utilization pools visits for Abdominal Pain, Gallbladder Disease, Bowel Obstruction, Constipation, Diarrhea, GERD, Gastritis, Gastroparesis, Nausea Vomiting, Noninfective Gastroenteritis, or Pancreatitis that were documented in any diagnosis position on medical claims after the index date, with a 30-day washout period before the index date to ensure that gastrointestinal symptoms were true side-effects rather than conditions present when the member initiated treatment. Utilization is assigned to the time period based on the start date of the stay or procedure.

[b] All costs were adjusted using the medical component of the Consumer Price Index for Urban Consumers to reflect inflation of costs between the time when medical care was provided and the end date of the current analysis (August 31, 2024). Costs were capped at $500,000 per year and scaled downward if the total medical and pharmacy costs exceeded that amount.

[c] Mean costs were compared with the Student's T Test. Without a Bonferroni correction for the 2 statistical tests done within each utilization category, population subgroup, and time period, a p-value less than 0.05 denotes a statistically significant difference, and a p-value less than 0.001 is a highly statistically significant difference. With a Bonferroni correction, only p-values less than 0.025 denote statistically significant differences, and p-values less than 0.0005 denote highly statistically significant differences.

[d] Inpatient medical admissions and inpatient surgical admissions were identified via diagnosis-related groupings (DRG).

**Table S7: Follow-up time of matched treated and control cohorts by quarter**

| | Full Population | | | | With Diabetes | | | | Without Diabetes | | | |
|---|---|---|---|---|---|---|---|---|---|---|---|---|
| | Treated | | Controls | | Treated | | Controls | | Treated | | Controls | |
| Quarters from Index | N | % Remaining | N | % Remaining | N | % Remaining | N | % Remaining | N | % Remaining | N | % Remaining |
| -4 | 742,824 | 100% | 742,824 | 100% | 413,404 | 100% | 413,404 | 100% | 329,420 | 100% | 329,420 | 100% |
| -3 | 742,824 | 100% | 742,824 | 100% | 413,404 | 100% | 413,404 | 100% | 329,420 | 100% | 329,420 | 100% |
| -2 | 742,824 | 100% | 742,824 | 100% | 413,404 | 100% | 413,404 | 100% | 329,420 | 100% | 329,420 | 100% |
| -1 | 742,824 | 100% | 742,824 | 100% | 413,404 | 100% | 413,404 | 100% | 329,420 | 100% | 329,420 | 100% |
| 0 (Year-Quarter of Index Date) | 742,824 | 100% | 742,824 | 100% | 413,404 | 100% | 413,404 | 100% | 329,420 | 100% | 329,420 | 100% |
| 1 | 742,824 | 100% | 742,824 | 100% | 413,404 | 100% | 413,404 | 100% | 329,420 | 100% | 329,420 | 100% |
| 2 | 708,095 | 95% | 704,400 | 95% | 396,139 | 96% | 392,839 | 95% | 311,956 | 95% | 311,561 | 95% |
| 3 | 624,288 | 84% | 606,250 | 82% | 347,695 | 84% | 336,989 | 82% | 276,593 | 84% | 269,261 | 82% |
| 4 | 547,400 | 74% | 516,421 | 70% | 302,374 | 73% | 285,804 | 69% | 245,026 | 74% | 230,617 | 70% |
| 5 | 443,962 | 60% | 414,960 | 56% | 251,886 | 61% | 235,084 | 57% | 192,076 | 58% | 179,876 | 55% |
| 6 | 339,310 | 46% | 318,823 | 43% | 205,757 | 50% | 190,995 | 46% | 133,553 | 41% | 127,828 | 39% |
| 7 | 275,589 | 37% | 252,590 | 34% | 175,739 | 43% | 158,383 | 38% | 99,850 | 30% | 94,207 | 29% |
| 8 | 229,016 | 31% | 207,960 | 28% | 150,893 | 37% | 134,093 | 32% | 78,123 | 24% | 73,867 | 22% |
| 9 | 189,936 | 26% | 170,989 | 23% | 129,269 | 31% | 113,279 | 27% | 60,667 | 18% | 57,710 | 18% |
| 10 | 157,303 | 21% | 140,399 | 19% | 110,114 | 27% | 95,342 | 23% | 47,189 | 14% | 45,057 | 14% |
| 11 | 133,795 | 18% | 117,027 | 16% | 95,520 | 23% | 80,849 | 20% | 38,275 | 12% | 36,178 | 11% |
| 12 | 113,528 | 15% | 98,302 | 13% | 82,448 | 20% | 68,824 | 17% | 31,080 | 9% | 29,478 | 9% |
| 13 | 96,694 | 13% | 83,020 | 11% | 71,232 | 17% | 58,739 | 14% | 25,462 | 8% | 24,281 | 7% |
| 14 | 82,568 | 11% | 70,516 | 9% | 61,080 | 15% | 49,961 | 12% | 21,488 | 7% | 20,555 | 6% |
| 15 | 71,503 | 10% | 60,682 | 8% | 52,901 | 13% | 43,005 | 10% | 18,602 | 6% | 17,677 | 5% |
| 16 | 62,106 | 8% | 52,231 | 7% | 46,283 | 11% | 37,099 | 9% | 15,823 | 5% | 15,132 | 5% |
| 17 | 54,501 | 7% | 45,320 | 6% | 40,832 | 10% | 32,243 | 8% | 13,669 | 4% | 13,077 | 4% |
| 18 | 46,789 | 6% | 38,625 | 5% | 35,068 | 8% | 27,472 | 7% | 11,721 | 4% | 11,153 | 3% |
| 19 | 39,985 | 5% | 32,685 | 4% | 29,936 | 7% | 23,151 | 6% | 10,049 | 3% | 9,534 | 3% |
| 20 | 33,776 | 5% | 27,450 | 4% | 25,387 | 6% | 19,356 | 5% | 8,389 | 3% | 8,094 | 2% |
| 21 | 28,244 | 4% | 22,919 | 3% | 21,257 | 5% | 16,086 | 4% | 6,987 | 2% | 6,833 | 2% |
| 22 | 23,160 | 3% | 18,905 | 3% | 17,365 | 4% | 13,197 | 3% | 5,795 | 2% | 5,708 | 2% |
| 23 | 19,070 | 3% | 15,491 | 2% | 14,282 | 3% | 10,754 | 3% | 4,788 | 1% | 4,737 | 1% |
| 24 | 15,449 | 2% | 12,536 | 2% | 11,540 | 3% | 8,696 | 2% | 3,909 | 1% | 3,840 | 1% |

**Figure S1. Total healthcare costs in quarter relative to initial prescribing of GLP-1RA: by GLP-1RA generation and diabetes status**

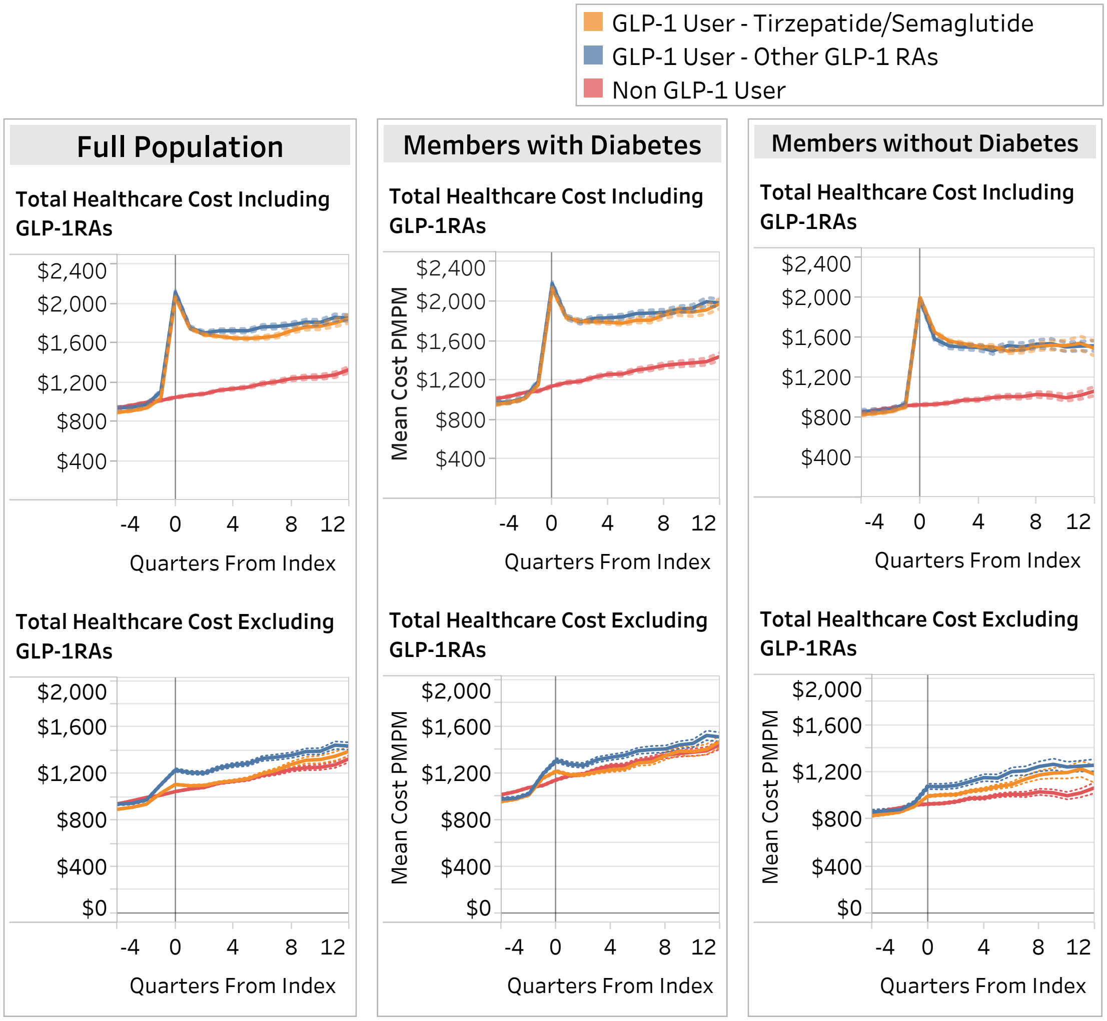


*Costs include both facility and professional claim types. Dotted lines are 95% confidence intervals.

**Figure S2. Costs* for select utilization categories in quarter relative to initial prescribing of GLP-1RA: by GLP-1RA generation and diabetes status**

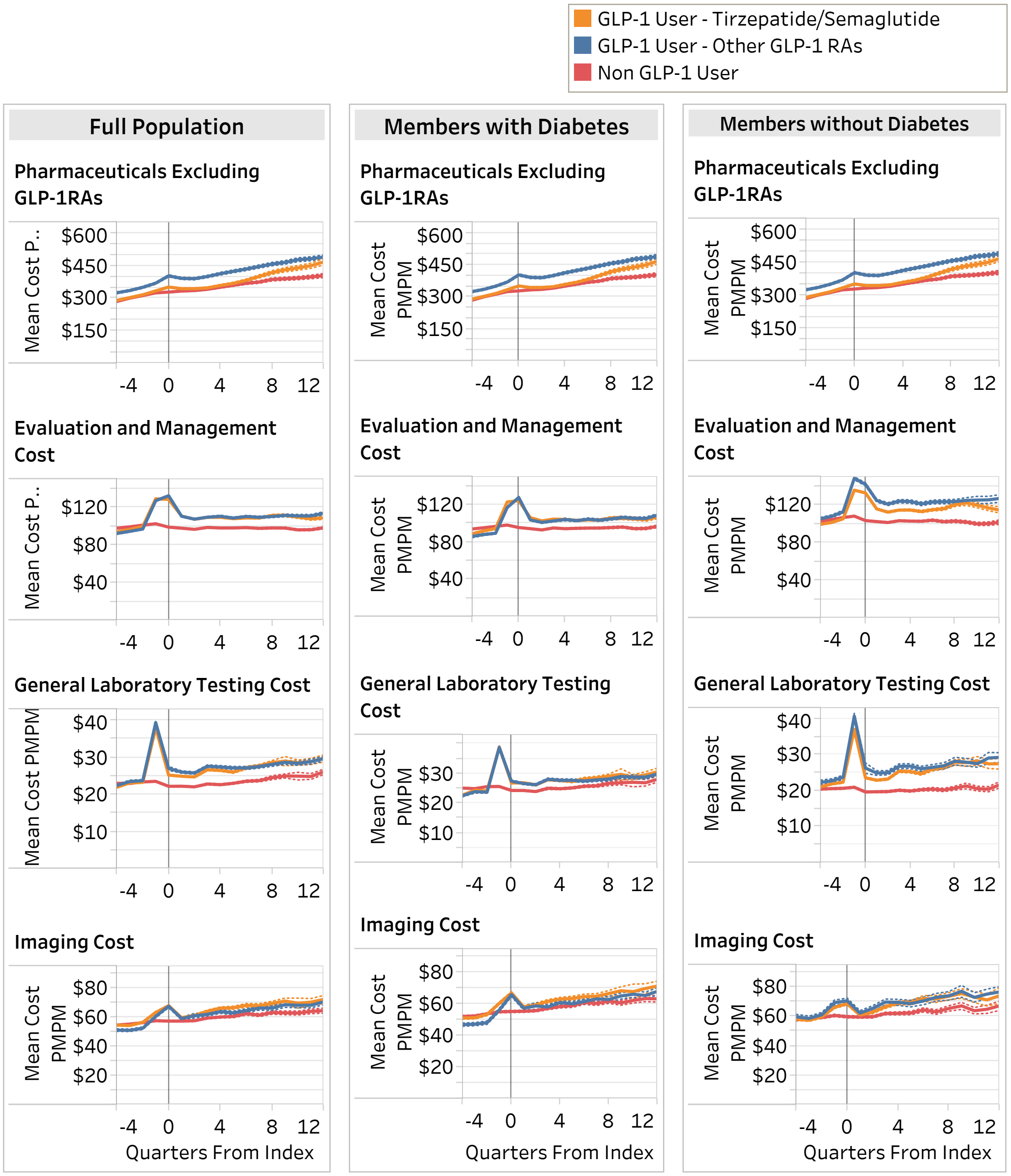


*Costs include both facility and professional claim types. Dotted lines are 95% confidence intervals.

**Figure S3. Inpatient medical admissions and dialysis costs* in quarter relative to initial prescribing of GLP-1RA: by GLP-1RA generation and diabetes status**

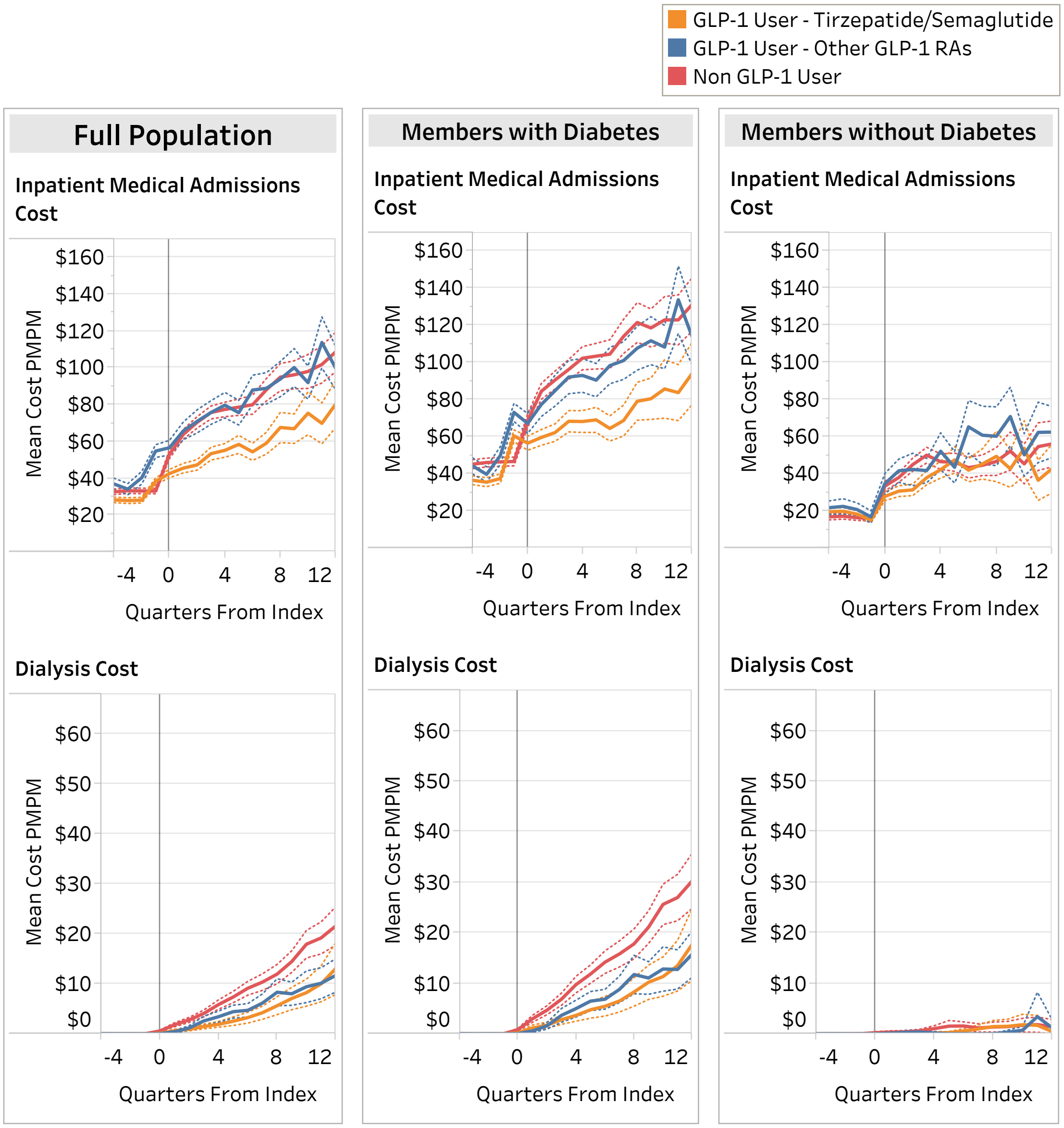


*Costs include both facility and professional claim types. Dotted lines are 95% confidence intervals.
